# Detection of RSV using nasopharyngeal swabs alone underestimates RSV-related hospitalization incidence in adults: the Multispecimen study’s Final Analysis

**DOI:** 10.1101/2025.01.14.25320406

**Authors:** Elizabeth Begier, Negar Aliabadi, Julio Ramirez, Allison McGeer, Qing Liu, Ruth Carrico, Samira Mubareka, Sonal Uppal, Stephen Furmanek, Zoe Zhong, Robin Hubler, Thomas Chandler, Caroline Kassee, Ashley Wilde, Kevin Katz, Paula Peyrani, Alan Junkins, Christie Vermeiren, Warren V Kalina, Ann R. Falsey, Edward Walsh, Malak Elsobky, Kari Yacisin, Elisa Gonzalez, Luis Jodar, Bradford D. Gessner

## Abstract

**Background:** RSV detection improves if an additional specimen is collected, but the impact of testing saliva and multiple specimen types has not been assessed. We quantified RSV detection increase with multiple specimen collection over nasopharyngeal swab (NPS) alone.

**Methods:** Prospectively enrolled hospitalized adults aged ≥40 years with acute respiratory illness in seven hospitals in US and Canada had NPS, saliva, sputa, and acute/convalescent sera collected and tested.

**Results:** Among 3,669 enrolled participants, 100% had NPS, 97.7% saliva, 33.0% sputum, and 33.4% paired serology. RSV detection was 112% higher (95% CI86%−141%) using all specimen types compared to NPS alone. Saliva had higher sensitivity than NPS (61.4% versus 47.2%). Among those with congestive heart failure exacerbations, additional specimens increased RSV detection by 267% (95% CI85%−625%) and saliva detected more infections than NPS. Among 1013 subjects with paired NPS from different timepoints tested on the same platform, specimens collected on average 1 day later detected 30% less RSV infections.

**Conclusions:** RSV detection increased over 100% using four specimen types versus NPS alone, suggesting a 2-fold correction factor is appropriate for incidence/prevalence studies relying on NPS alone. Saliva is more sensitive than NPS, warranting further study particularly in cardiac patients.

## INTRODUCTION

Respiratory syncytial virus (RSV) is increasingly recognized as an important cause of respiratory tract illness among older adults [1, 2] and adults with immunocompromise or other underlying chronic co-morbidities [3–5]. Pooled analyses of prospective studies have reported that, after adjustment for diagnostic testing-based under ascertainment, the annual RSV-related hospitalization incidence rate in adults 65 years of age and older is 282 per 100,000 [6] in the US and 347 per 100,000 in high income countries [1]. Most published RSV incidence rates are recognized as underestimates for several reasons [7]. First, RSV testing is infrequently conducted as part of standard of care (SOC) [8], partly owing to lack of RSV-specific treatment. Second, case definitions do not always include the full clinical spectrum of RSV disease (e.g., case definitions using pneumonia only or requiring fever) [9, 10]. Third, many studies use limited sampling periods (e.g. period of peak influenza activity that is not always aligned with the RSV activity period) [11]. Fourth, diagnostic challenges, including delays between illness onset and specimen sampling, low viral load [12], and reliance on a single obtained specimen, may lead to missed infections [13, 14].

Although nasopharyngeal swabs (NPS) are the most commonly used specimens for diagnostic testing of respiratory viruses [6], sputum and other lower respiratory tract specimens (e.g., bronchoalveolar lavage [BAL], endotracheal aspirates) have higher sensitivity for RSV in older adults [15, 16] who may shed less virus in nasal secretions as compared to children [12, 17, 18]. A recent systematic literature review (SLR) and meta-analysis found that adding a single specimen to nasal/NPS reverse transcriptase (RT)-polymerase chain reaction (PCR) testing increased RSV detection by 52% with sputum, 44% with paired serology, and 28% with oropharyngeal swabs [13].

Previously, we reported on a preliminary analysis of 1.5 seasons of US data from the present study [14]. In this report, we expanded on the original cohort of 1766 US participants enrolled by adding complete results from an additional season and including Canadian sites, thus doubling the number of participants. The primary objective was to quantify the underestimation in RSV diagnosis associated with using NPS RT-PCR alone by comparing the yield of NPS alone to the yield by combinations of additional specimen types, overall and by geographic setting. Additional analyses explore whether participant characteristics affect diagnostic accuracy of specimens, and comparison of RSV detection between salvaged standard-of-care (SOC) and study NPS specimens.

## METHODS

This prospective cohort study enrolled adults in four hospitals in Louisville, Kentucky over 2 RSV seasons, from 27Dec2021−01Apr2022, and 22Aug2022−03Mar2023 (WCG IRB approval #21-N0325) and three hospitals in Ontario, Canada (IRB approvals # 22-0096-E, #5522 and #2023-0197-562) over 1 RSV season from 12Oct2022−03Mar2023. Informed consent was obtained from all patients. Methods have been previously described [14]. Hospitals in Louisville provided general adult acute care and those in Ontario included 2 tertiary care hospitals and one community teaching hospital. In both settings, patients aged ≥40 years who were hospitalized with acute respiratory illness (ARI) or ARI-compatible symptoms with onset <21 days prior to admission and who were willing to provide an NPS and at least one other diagnostic specimen were enrolled (Supplementary Table 1 provides full eligibility criteria). Enrollment was through active surveillance during the RSV season, which was defined by local RSV surveillance.

ARI or ARI-compatible symptoms were defined as either: 1) new onset or increase from baseline of: nasal congestion, rhinorrhea, sore throat, hoarseness, cough, sputum production, dyspnea, wheezing, hypoxemia, or 2) admitting diagnosis suggestive of ARI (e.g., pneumonia, upper respiratory infection, bronchitis, influenza, cough, viral respiratory illness, respiratory distress, or respiratory failure), or 3) admitting diagnosis was the exacerbation of an underlying cardiac or pulmonary disease involving acute respiratory symptoms (e.g., congestive heart failure [CHF], chronic obstructive pulmonary disease [COPD], or asthma exacerbation). Patients were excluded if they developed ARI symptoms ≥48 hours after admission, or if previously enrolled in the study in the last 45 days. After consent, specimens and clinical, demographic, social, medical and vaccination information were collected from medical records and patient questionnaires. At a second study visit scheduled for 30−60 days after enrolment, information regarding clinical course, intercurrent ARIs, and convalescent serum was collected.

### Study specimen collection

At enrollment, study NPS were obtained from all participants, plus at least one of the following study specimens: saliva, sputum, acute serum. Every effort was made to collect all additional specimen types in as short a period of time as possible (e.g. ideally within 24 hours of hospitalization). If these specimen types were available from previously collected SOC specimens not tested for viral RNA, they were salvaged for study testing. Patients unable to produce saliva provided a saline swish and spit using 2mL of normal saline at US sites and 3mL at Canadian sites. SOC saliva specimens were not available at either site. If no study sputum specimen could be obtained, the salvaged sputum specimen collected closest in time and within 36 hours of the study NP (if available) was included as a study specimen in the primary analysis. At the second study visit, convalescent serum was collected and NPS for subjects reporting intercurrent ARI. Additionally, at the Toronto site, NPS were also collected at ARI onset between visits when feasible.

### RSV Diagnosis from study specimens

At the US sites, respiratory specimens were tested for RSV by RT-PCR using ARIES Luminex FluA/B/RSV panel (Luminex), which is approved for use on NPS with 97.1% positive agreement and 98.4% negative agreement with an FDA cleared comparator [19]. At the Canadian sites, all study samples were tested using a local laboratory developed RT-PCR respiratory virus panel (LDT-RP) targeting RSV A/B nucleocapsid (N) gene, which was validated for NPS, sputum/(BAL) and saliva. Briefly, samples were extracted using the Maxwell Viral HT TNA kit (Promega) and amplified using Luna Universal Probe One-Step RT-qPCR Kit (New England Biolabs). Study and SOC samples negative for RSV or positive with a cycle threshold (Ct of >30) when tested by LDT-RP, and which had sufficient volume remaining for testing, had repeat testing using the Luminex to confirm results between assay types, with 96% agreement. Any specimen identified as positive by either LDT-RP or Luminex was considered positive in this analysis. Acute and convalescent sera were tested for levels of antibodies against non-vaccine RSV antigens (matrix, nucleoprotein, and G [Ga or Gb peptides]) in a Luminex-based immunoassay at the Pfizer Vaccine Research and Development central lab. The sources of recombinant matrix protein, nucleoprotein and G protein peptide sequences (Cambridge Research Biochemicals) were previously described [14, 20]. A four-fold rise between acute and convalescent paired sera for any of these four antigens indicated RSV infection.

### Analyses

Patients were included if they had results from a study NPS and at least one other study specimen. Serology results were included only if results from both acute and convalescent specimens were available.

RSV prevalence was defined as the percentage of participants with at least one RSV positive study specimen and was generated for study NPS alone, and for study NPS in various combinations with study sputum, serology, or saliva. The percent increase in RSV detection was calculated by the following formula:

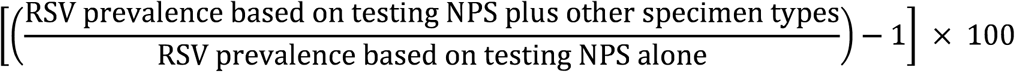

with 95% confidence intervals (CIs) calculated. Estimates were calculated for the study population overall and limited to those with the specific combinations of specimen type results available (e.g., had both sputum and NPS results for that specific percent increase). Estimates were similarly generated for subpopulations of patients with different characteristics. Sensitivity was calculated by specimen type, limited to those with results with that specimen type, calculated as the count of positives by that specimen type divided by count of all positives from any source among those with results by that specimen type. A composite reference for defining true positivity was defined as positive by any specimen type and was used to calculate sensitivity [13].

Among patients with multiple NPS collected for their ARI hospitalization, with RSV testing conducted on study assays, we described the time to the first and second NPS and compared the number of RSV detections in the first NPS to that in the second NPS.

## RESULTS

Overall, 3,669 participants met eligibility criteria, were enrolled, had a valid study NPS and at least one other study specimen tested and were thus included in the primary analysis (Figure 1).

**Figure 1.**
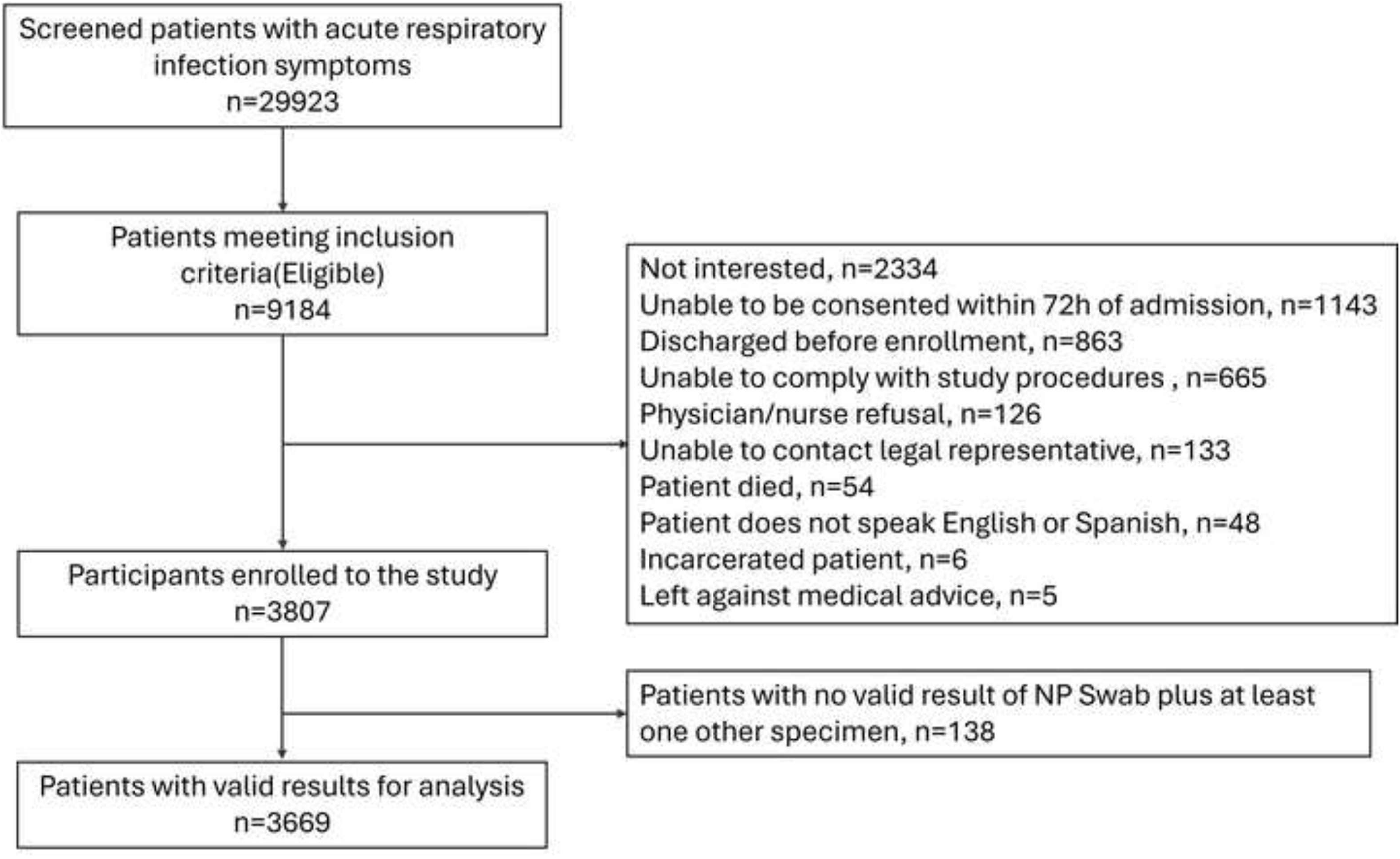
Enrolment flow diagram, US and Canadian sites NP, nasopharyngeal

### Patient characteristics

The majority (76%, 2790/3669) of patients were from the US site. Overall, median age was 68 years (IQR 58−77); 45.1% were male; and 68.2% were White, 19.7% Black and 2.3% Hispanic (Table 1). Canadian patients were older than US patients (median age: 78 versus 65 years). A higher proportion of Canadian patients were Asian and lower proportion Black compared to the US participants. Baseline comorbidities were common overall including chronic heart disease (n=1490, 40.6%), diabetes mellitus (n=1349, 36.8%), COPD (n=1264, 34.5%), CHF (n=1143, 31.2%), and coronary artery disease (CAD; n=1100, 30.0%), with higher proportions of US vs. Canadian patients having these conditions. Disease severity as measured by ICU stay or use of mechanical ventilation was 9.5% overall and similar between sites; however, a higher proportion of Canadian patients died during hospitalization (2.9% overall, 5.9% Canada vs 1.9% US, Supplementary Table 2).

**Table 1:** Selected characteristics of study participants.

|  | **US** |  | **Canada** |  | **All sites** |  | **P Value<sup>a</sup>** |
| --- | --- | --- | --- | --- | --- | --- | --- |
|  | **N** | **%** | **N** | **%** | **N** | **%** |  |
| Total enrolled | 2790 | 100 | 879 | 100 | 3669 | 100 |  |
| Demographics |  |  |  |  |  |  |  |
| Age in Years |  |  |  |  |  |  |  |
| Median (IQR) | 65 | 56.0–<br>74.0 | 78 | 68.0–<br>87.0 | 68 | 58.0–<br>77.0 | <.0001 |
| 40-49 | 376 | 13.5 | 27 | 3.1 | 403 | 11.0 | <.0001 |
| 50-59 | 572 | 20.5 | 73 | 8.3 | 645 | 17.6 |  |
| 60-64 | 418 | 15.0 | 63 | 7.2 | 481 | 13.1 |  |
| 65-74 | 753 | 27.0 | 180 | 20.5 | 933 | 25.4 |  |
| 75-84 | 509 | 18.2 | 251 | 28.6 | 760 | 20.7 |  |
| ≥85 | 162 | 5.8 | 285 | 32.4 | 447 | 12.2 |  |
| Gender |  |  |  |  |  |  |  |
| Male | 1196 | 42.9 | 458 | 52.1 | 1654 | 45.1 | <.0001 |
| Race<sup>b</sup> |  |  |  |  |  |  |  |
| White | 1961 | 70.3 | 541 | 61.4 | 2502 | 68.2 | N/A |
| Black | 694 | 24.9 | 30 | 3.4 | 724 | 19.7 |  |
| Southeast or South Asian | 14 | 0.5 | 253 | 28.7 | 267 | 7.3 |  |
| Indigenous/First Nations | 4 | 0.1 | 3 | 0.3 | 7 | 0.2 |  |
| Other | 115 | 4.1 | 18 | 2.0 | 133 | 3.6 |  |
| Latin American | 0 | 0 | 13 | 1.5 | 13 | 0.4 |  |
| Undefined | 2 | 0.1 | 23 | 2.6 | 25 | 0.7 |  |
| Ethnicity (US only)<sup>c</sup> |  |  |  |  |  |  |  |
| Hispanic or Latino | 83 | 3.0 | 0 | 0 | 83 | 2.3 | N/A |
| Not Hispanic or Latino | 2707 | 97.0 | 0 | 0 | 2707 | 73.8 |  |
| Undefined | 0 | 0 | 879 | 100 | 879 | 24.0 |  |
| Substance Use |  |  |  |  |  |  |  |
| Tobacco |  |  |  |  |  |  |  |
| Yes | 1263 | 45.3 | 255 | 29.0 | 1518 | 41.4 | <0.0001 |
| Comorbidities |  |  |  |  |  |  |  |
| Chronic heart disease, other | 1149 | 41.2 | 341 | 38.8 | 1490 | 40.6 | 0.2086 |
| Diabetes mellitus | 1090 | 39.1 | 259 | 29.5 | 1349 | 36.8 | <.0001 |
| Chronic obstructive pulmonary disease | 1087 | 39 | 177 | 20.1 | 1264 | 34.5 | <.0001 |
| Congestive heart failure | 932 | 33.4 | 211 | 24.0 | 1143 | 31.2 | <.0001 |
| Coronary artery disease | 907 | 32.5 | 193 | 22.0 | 1100 | 30.0 | <.0001 |
| Chronic kidney disease | 658 | 23.6 | 176 | 20.0 | 834 | 22.7 | 0.028 |
| Asthma | 563 | 20.2 | 108 | 12.3 | 671 | 18.3 | <.0001 |
| Cancer, solid tumor | 545 | 19.5 | 206 | 23.4 | 751 | 20.5 | 0.0124 |
| Neurologic and neurodevelopment conditions | 499 | 17.9 | 168 | 19.1 | 667 | 18.2 | 0.4107 |
| Stroke | 352 | 12.6 | 89 | 10.1 | 441 | 12.0 | 0.0476 |
| Chronic lung disease, other | 255 | 9.1 | 86 | 9.8 | 341 | 9.3 | 0.5663 |
| Immunosuppressant drug therapy (active) | 240 | 8.6 | 119 | 13.5 | 359 | 9.8 | <.0001 |
| Autoimmune disorders | 199 | 7.1 | 47 | 5.3 | 246 | 6.7 | 0.0649 |
| Chronic liver disease | 139 | 5 | 30 | 3.4 | 169 | 4.6 | 0.053 |
| Cancer, hematologic malignancy | 93 | 3.3 | 84 | 9.6 | 177 | 4.8 | <.0001 |
| Blood disorder | 68 | 2.4 | 21 | 2.4 | 89 | 2.4 | 0.9354 |
| HIV | 36 | 1.3 | 2 | 0.2 | 38 | 1.0 | 0.0066 |
| Organ transplant | 17 | 0.6 | 10 | 1.1 | 27 | 0.7 | 0.11 |
| Immunodeficiency | 11 | 0.4 | 95 | 10.8 | 106 | 2.9 | <.0001 |
| No comorbidities | 175 | 6.3 | 76 | 8.6 | 251 | 6.8 | 0.0151 |
| Risk Classification |  |  |  |  |  |  |  |
| High | 2615 | 93.7 | 803 | 91.4 | 3418 | 93.2 | 0.0151 |
| Immunocompromised | 839 | 30.1 | 342 | 38.9 | 1181 | 32.2 | <.0001 |
| Not immunocompromised | 1776 | 63.7 | 461 | 52.4 | 2237 | 61.0 |  |
| Low | 175 | 6.3 | 76 | 8.6 | 251 | 6.8 |  |
| Disease Severity |  |  |  |  |  |  |  |
| ICU | 247 | 8.9 | 86 | 9.8 | 333 | 9.1 | 0.4039 |
| Mechanical ventilation | 79 | 2.8 | 29 | 3.3 | 108 | 2.9 | 0.4796 |
| ICU or mechanical ventilation | 262 | 9.4 | 86 | 9.8 | 348 | 9.5 | 0.7287 |
| Not severe | 2528 | 90.6 | 793 | 90.2 | 3321 | 90.5 |  |

N, number of patients; IQR, interquartile range
<sup>a</sup>P value compares participants in the US to those in Canada
<sup>b</sup>Participants from Canadian sites may report multiple races.
<sup>c</sup>Canadian definitions do not distinguish race and ethnicity, and do not include Hispanic/Latino as a category.

### Study specimens and RSV prevalence

Overall, 254 (6.9%) persons tested positive for RSV in at least one study sample type: 86 (9.8%) at Canadian sites and 168 (6.0%) from US sites (Table 2). Among 3,669 patients enrolled with NPS collected, 3583 (97.7%) provided saliva or saline mouth wash, 1212 (33.0%) provided sputum, and 1225 (33.4%) provided paired sera. Of those providing saliva, 541 (19.7%) in US and 267 (32.0%) in Canada provide saline mouth wash rather than neat saliva (Supplementary Table 3). RSV was detected in 3.3% of NPS (2.8% US vs 4.8% Canada), 4.3% of saliva samples (3.9% US vs 5.6% Canada), 7.3% of sputum samples (6.1% US vs 10.7% Canada), and 7.3% of paired serology samples (7.2% US vs 7.3% Canada). Saliva specimens identified the highest absolute number of positive RSV infections (n=153). All specimen types added unique positive RSV detections (Figure 2). Among 265 participants who reported an intercurrent ARI at or before visit 2 and with NPS tested, 6 were RSV positive (2 from US and 6 from Canadian sites).

**Figure 2.**
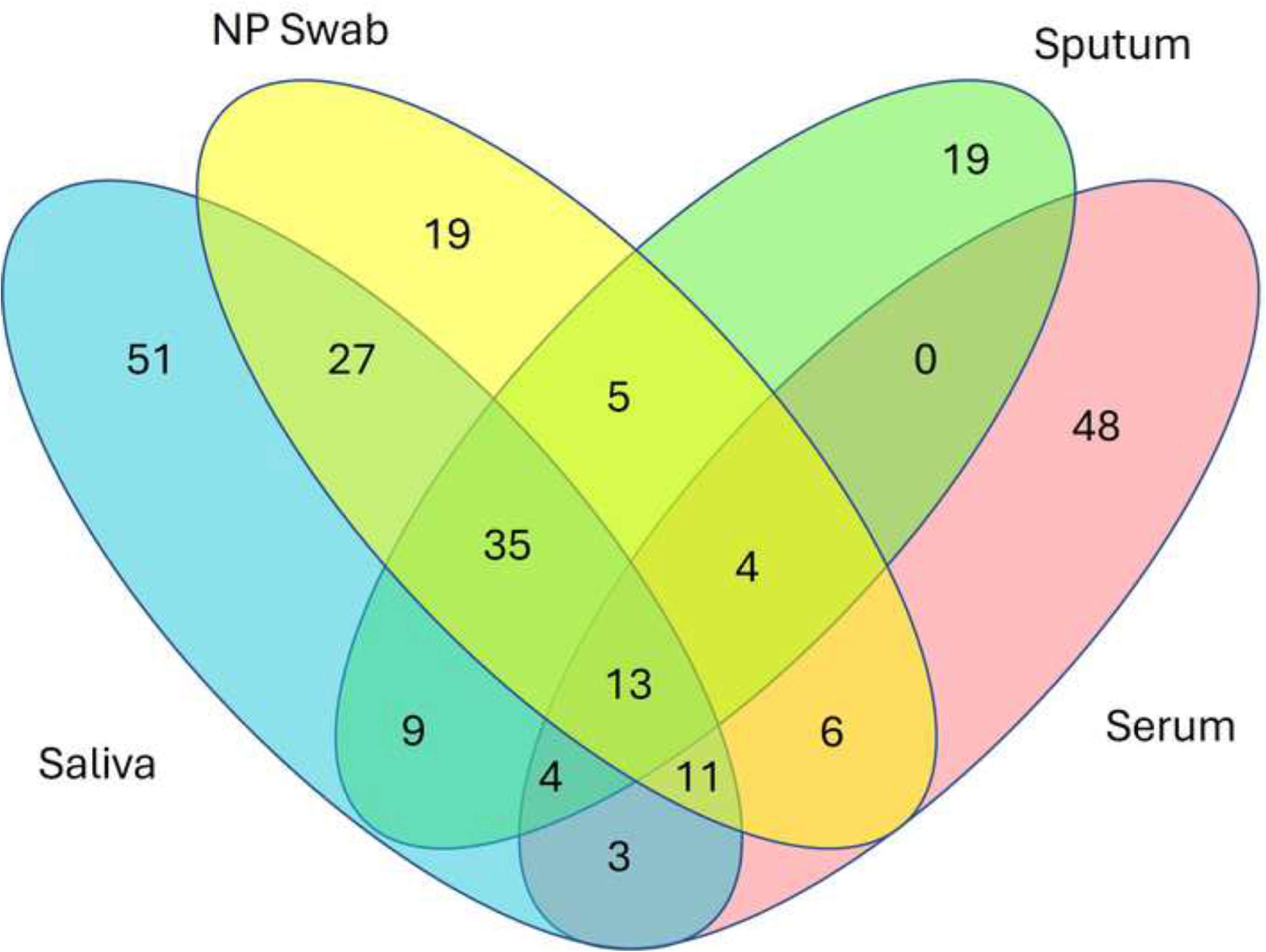
RSV diagnosis by combinations of specimen types: nasopharyngeal swab, saliva, sputum and serology NP, nasopharyngeal

**Table 2:** Specimen collection and RSV test results for study specific testing.

|  | US |  | Canada |  | All Sites |  |
| --- | --- | --- | --- | --- | --- | --- |
| **Study Specimen Collection** | **N** | **%** | **N** | **%** | **N** | **%** |
| Total enrolled | 2790 | 100 | 879 | 100 | 3669 | 100 |
| Participants with any RSV positive result | 168 | 6.0 | 86 | 9.8 | 254 | 6.9 |
| Study Specimen Test Results<sup>a</sup> |  |  |  |  |  |  |
| NPS | 2790 |  | 879 |  | 3669 |  |
| Positive | 78 | 2.8 | 42 | 4.8 | 120 | 3.3 |
| Negative | 2712 | 97.2 | 837 | 95.2 | 3549 | 96.7 |
| Saliva/saline wash | 2749 |  | 834 |  | 3583 |  |
| Positive | 106 | 3.9 | 47 | 5.6 | 153 | 4.3 |
| Negative | 2643 | 96.1 | 787 | 94.4 | 3430 | 95.7 |
| Sputum | 874 |  | 338 |  | 1212 |  |
| Positive | 53 | 6.1 | 36 | 10.7 | 89 | 7.3 |
| Negative | 821 | 93.9 | 302 | 89.3 | 1123 | 92.7 |
| Serum<sup>b</sup> | 843 |  | 382 |  | 1225 |  |
| 4-fold rise | 61 | 7.2 | 28 | 7.3 | 89 | 7.3 |
| 2-fold rise | 129 | 15.3 | 58 | 15.2 | 187 | 15.3 |
| Negative | 714 | 84.7 | 324 | 84.8 | 1038 | 84.7 |

N, number of patients
<sup>a</sup>Includes study testing of salvaged specimens
<sup>b</sup>Serum is only counted if paired acute and convalescent specimens are collected.

Most study participants had an SOC specimen tested for RSV, and some had other respiratory specimens tested as SOC or salvaged for study testing (Supplementary Table 4). These specimens were more frequently RSV positive: 6.3% (157/2394) NPS, 18.5% (5/27) sputum and 6.1% (2/33) BAL. The 2 participants with RSV-positive BAL specimens also had RSV detected in multiple other specimen types. Small numbers of throat swabs, tracheal aspirates, pleural fluid specimens, nasal washes, nasal swabs, and oropharyngeal swabs were obtained: none yielded RSV.

### Sensitivity

We examined each specimen type’s sensitivity by limiting analysis to participants with that result type available (Table 3), using as the gold standard positivity by any specimen type. Serology sensitivity was the highest (73.0%; 95% CI 65.1−80.8), followed by sputum (70.1%; 95%CI 62.1−78.0), saliva (61.4%; 95%CI 55.4−67.5), and NPS (47.2%; 95%CI 41.1−53.4).

**Table 3:** Specimen collection, RSV prevalence and sensitivity by specimen type.

| Study specimens | N | Percentage of participants with this specimen type | n, positive by specimen type | %, positive by specimen type | Percentage of all positives identified by this specimen type | 95% CI | RSV+ from any specimen among those with that specimen type | Sensitivity | 95% CI |
| --- | --- | --- | --- | --- | --- | --- | --- | --- | --- |
| Any | 3669 | 100% | 254 | 6.9% | N/A | N/A | 254 | N/A | N/A |
| NP swab | 3669 | 100% | 120 | 3.3% | 47.2% | 41.1–53.4% | 254 | 47.2% | 41.1–53.4% |
| Saliva<sup>b</sup> | 3583 | 97.7% | 153 | 4.3% | 60.2% | 54.2–66.3% | 249 | 61.4% | 55.4–67.5% |
| Sputum | 1212 | 33.0% | 89 | 7.3% | 35.0% | 29.2–40.9% | 127 | 70.1% | 62.1–78.0% |
| Serum | 1225 | 33.4% | 89 | 7.3% | 35.0% | 29.2–40.9% | 122 | 73.0% | 65.1–80.8% |

N, total number of patients; n, number of patients with a RSV-positive specimen
<sup>a</sup>Sensitivity is limited to those with results from that specimen type, calculated as the number of RSV positive specimens for a particular type divided by number of all RSV positive specimens from any source among those with results by that specimen type
<sup>b</sup>In subjects that were unable to produce saliva, a saline mouth wash was obtained.

### RSV detection

Among all enrolled participants regardless of specific specimens available, 254 (6.9%) had RSV diagnosed by any specimen versus 120 (3.3%) diagnosed by NPS alone, representing a 112% increase in RSV detection (95%CI 86−141%). A stepwise increase in RSV detection was seen with the addition of specimen types (Figure 3A; Supplementary Table 5). Among 417 patients with all four specimen types collected, RSV detection was 117% (95%CI 62−191%) higher compared to those with NPS alone (Figure 3B; Supplementary Table 6). Limiting to those with NPS and the specific second specimen type available, RSV detection increased 115% (95%CI 75−164%) with serum, 57% (95%CI 41−75%) with saliva, and 48% (95%CI 29−70%) with sputum.

**Figure 3.**
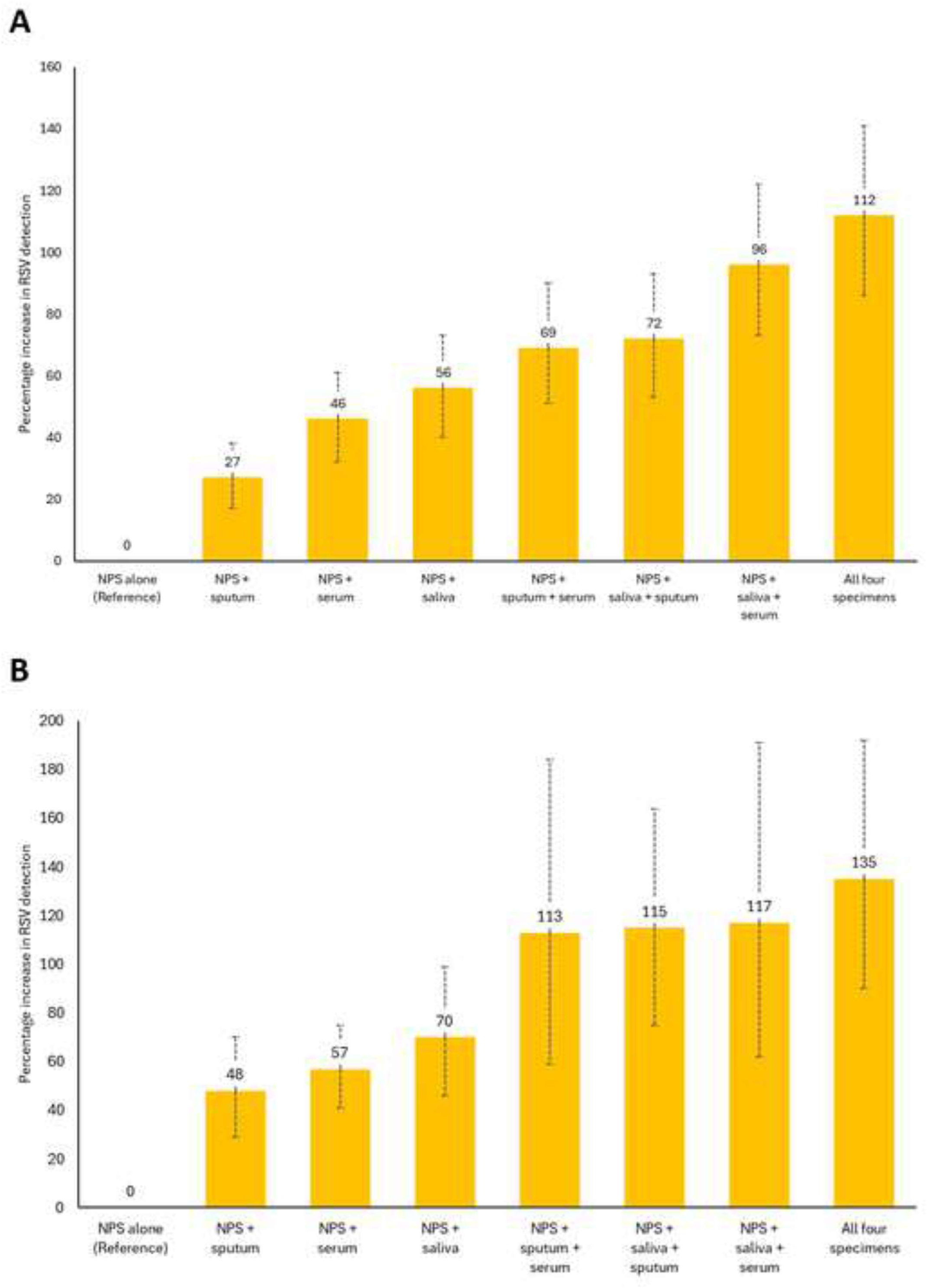
The percent of RSV diagnoses increases when adding specimen types over using NPS alone Panel A: For all participants in the study (see Supplemental Table 5) Panel B: For different populations with specific specimen results available (see Supplemental Table 6) NPS, nasopharyngeal swab

We conducted subgroup analyses of the percent increase in RSV detection comparing multiple specimens to NPS alone among the entire study population (Table 4). Overall, the percent increase in RSV detection was greater among those aged 40−64 years (141%, 95%CI 92−202%) than among those >65 years (95%, 95%CI 66−128%). Compared to NPS alone, RSV detection by multiple specimens ranged from 88−200% higher at all timepoints from onset of symptom to testing. Additionally, there was a higher RSV detection by multiple specimens compared to NPS alone for both immunocompromised and immunocompetent individuals, for all individual signs and symptoms except hoarseness, and for all final clinical diagnoses except empyema which did not have enough observations to generate an estimate. Of note, participants admitted with ARI who had final diagnoses of common cardiac conditions had the highest increase in RSV detection: CHF 267% (95%CI 85−625%); acute chest pain/cardiac ischemia 700% (95%CI 28−4904%), although confidence intervals were wide.

**Table 4:**
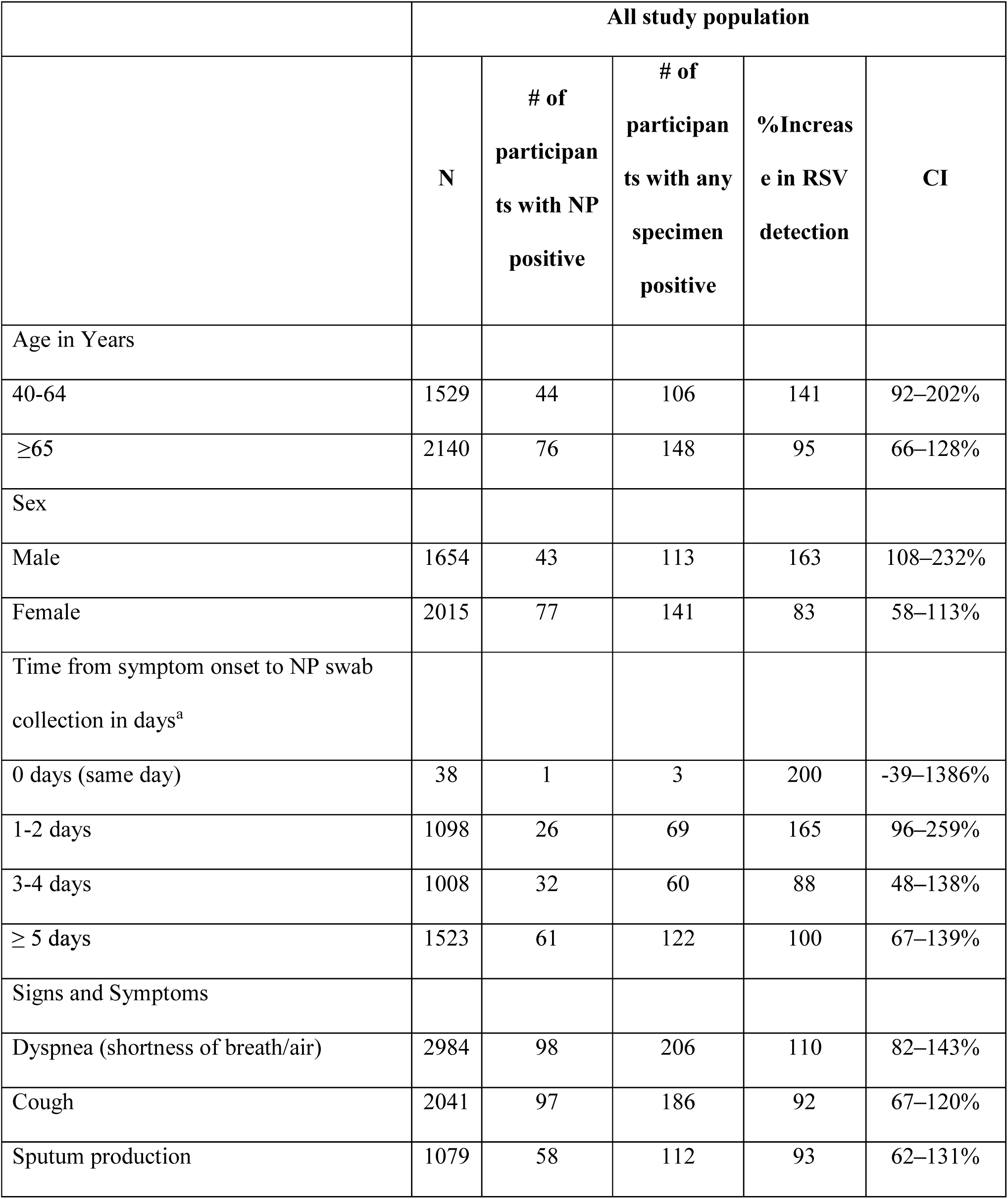
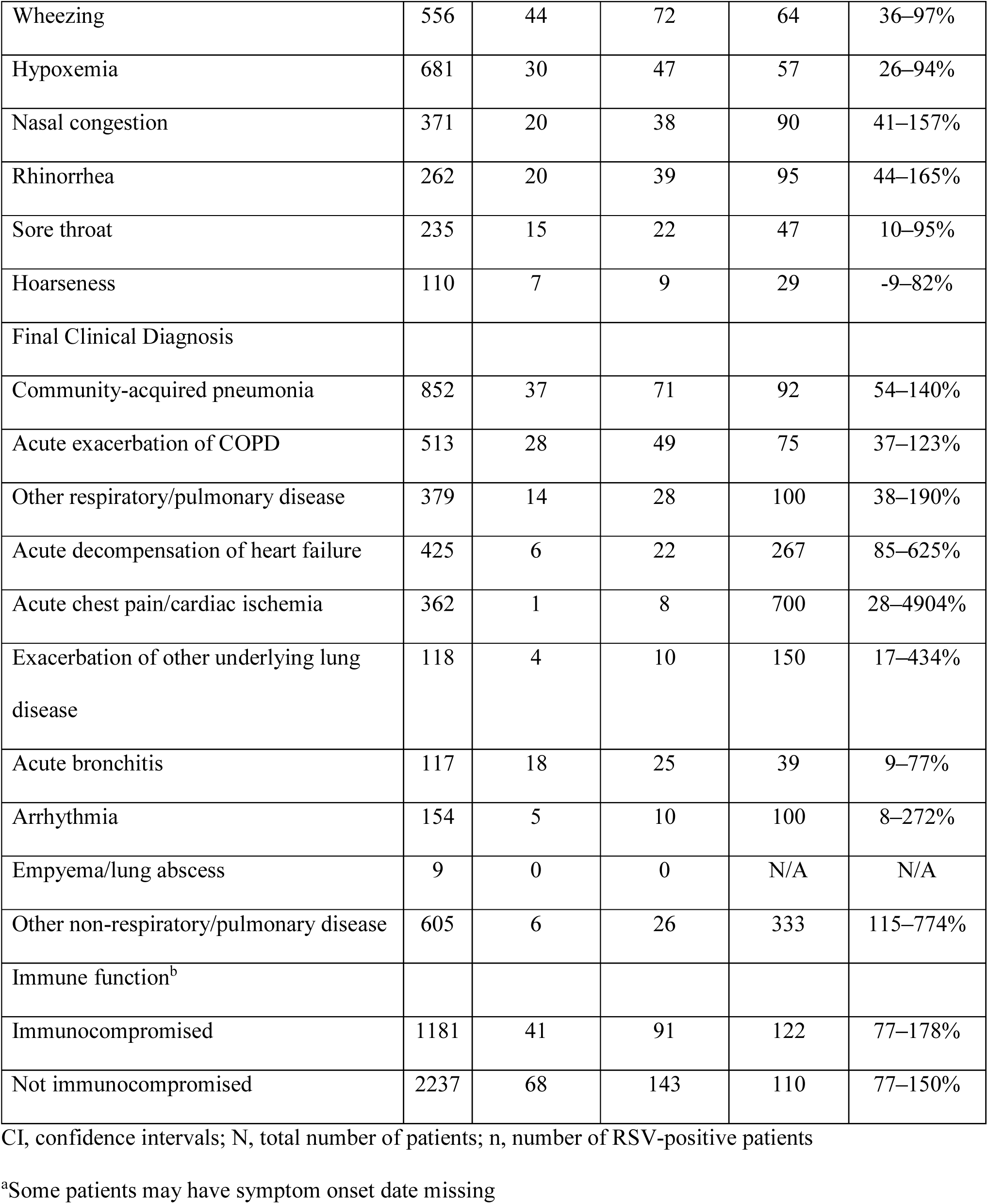
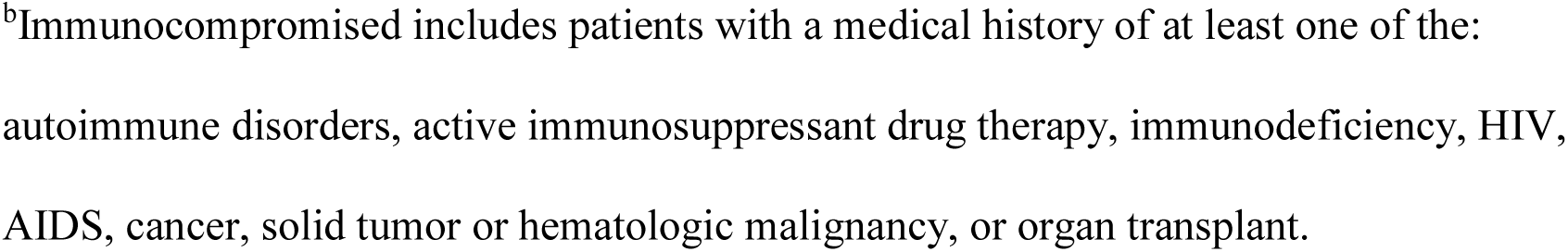
Increase in RSV detection associated with multiple specimen use for all participants in the study, by participant characteristics.

|  | All study population |  |  |  |  |
| --- | --- | --- | --- | --- | --- |
|  | N | # of participants with NP positive | # of participants with any specimen positive | %Increase in RSV detection | CI |
| Age in Years |  |  |  |  |  |
| 40-64 | 1529 | 44 | 106 | 141 | 92–202% |
| ≥65 | 2140 | 76 | 148 | 95 | 66–128% |
| Sex |  |  |  |  |  |
| Male | 1654 | 43 | 113 | 163 | 108–232% |
| Female | 2015 | 77 | 141 | 83 | 58–113% |
| Time from symptom onset to NP swab collection in days<sup>a</sup> |  |  |  |  |  |
| 0 days (same day) | 38 | 1 | 3 | 200 | -39–1386% |
| 1-2 days | 1098 | 26 | 69 | 165 | 96–259% |
| 3-4 days | 1008 | 32 | 60 | 88 | 48–138% |
| ≥ 5 days | 1523 | 61 | 122 | 100 | 67–139% |
| Signs and Symptoms |  |  |  |  |  |
| Dyspnea (shortness of breath/air) | 2984 | 98 | 206 | 110 | 82–143% |
| Cough | 2041 | 97 | 186 | 92 | 67–120% |
| Sputum production | 1079 | 58 | 112 | 93 | 62–131% |
| Wheezing | 556 | 44 | 72 | 64 | 36–97% |
| Hypoxemia | 681 | 30 | 47 | 57 | 26–94% |
| Nasal congestion | 371 | 20 | 38 | 90 | 41–157% |
| Rhinorrhea | 262 | 20 | 39 | 95 | 44–165% |
| Sore throat | 235 | 15 | 22 | 47 | 10–95% |
| Hoarseness | 110 | 7 | 9 | 29 | -9–82% |
| Final Clinical Diagnosis |  |  |  |  |  |
| Community-acquired pneumonia | 852 | 37 | 71 | 92 | 54–140% |
| Acute exacerbation of COPD | 513 | 28 | 49 | 75 | 37–123% |
| Other respiratory/pulmonary disease | 379 | 14 | 28 | 100 | 38–190% |
| Acute decompensation of heart failure | 425 | 6 | 22 | 267 | 85–625% |
| Acute chest pain/cardiac ischemia | 362 | 1 | 8 | 700 | 28–4904% |
| Exacerbation of other underlying lung disease | 118 | 4 | 10 | 150 | 17–434% |
| Acute bronchitis | 117 | 18 | 25 | 39 | 9–77% |
| Arrhythmia | 154 | 5 | 10 | 100 | 8–272% |
| Empyema/lung abscess | 9 | 0 | 0 | N/A | N/A |
| Other non-respiratory/pulmonary disease | 605 | 6 | 26 | 333 | 115–774% |
| Immune function<sup>b</sup> |  |  |  |  |  |
| Immunocompromised | 1181 | 41 | 91 | 122 | 77–178% |
| Not immunocompromised | 2237 | 68 | 143 | 110 | 77–150% |

CI, confidence intervals; N, total number of patients; n, number of RSV-positive patients
<sup>a</sup>Some patients may have symptom onset date missing
<sup>b</sup>Immunocompromised includes patients with a medical history of at least one of the: autoimmune disorders, active immunosuppressant drug therapy, immunodeficiency, HIV, AIDS, cancer, solid tumor or hematologic malignancy, or organ transplant.

The degree of additional infections detected by using multiple specimens varied by patient characteristics (Supplementary table 7). Median time from symptom onset to specimen collection was 3.0−4.0 days among RSV+ events identified by respiratory specimens compared to 5.0 days among those identified by serology. Among those with RSV detections by saliva (saliva+/NPS-), diagnoses of acute CHF and acute chest pain/cardiac ischemia were more frequent (11.9% and 3%, respectively) compared to NPS positive participants (5% and 0.8%).

### Comparison of paired NPS

Among a subset of 1013 participants with paired NPS from different timepoints tested on the same platform, 56 positives were identified from the chronologically first NPS and only 39 of these 56 also had RSV detected in their second NPS. Median time from symptom onset to the first NPS was 3.0 days (IQR 1.0−6.0 days) compared to 4.0 days (IQR 2.0−7.0 days) for the second NPS. Thus, the later NPS detected only 70% of RSV infections compared to the NPS that was collected only 1 day earlier on average equating to a 30% decrease in detection with 1 day delay.

## DISCUSSION

In this full analysis, we found the addition of multiple specimen types – compared to the current research and clinical standard practice of single NPS alone – in patients being hospitalized with respiratory illness approximately doubles RSV detection. The increased detection with multiple specimens was seen across geographically, demographically, and clinically heterogeneous populations. A trend towards higher detection was seen among participants with cardiac diagnoses, particularly with saliva testing. The data also highlight saliva testing as a sensitive tool and the importance of NPS specimen collection timing. Saliva/saline mouthwash testing identified the most positives due to collection for nearly all participants and higher sensitivity than NPS. Among participants with paired NPS specimens, those collected an average 1 day later than the first specimens detected 30% less RSV. These findings add to the growing literature indicating single specimen testing and delayed specimen collection with NPS underestimates RSV infection among adults by about half.

Overall, in the entire study population, we found a 112% higher RSV detection rate when additional specimens were obtained beyond NPS, an increase that occurred to varying degrees among adults with differing baseline characteristics. The addition of collection of one specimen type (saliva, sputum, or paired serology specimens) to NPS alone increased RSV detection by 56%, 27%, and 46%, respectively. Notably, adding collection of both saliva and sputum, samples which may be available during acute illness, increased yield 72%. Our data suggest that studies based on NPS alone should use an approximate 2-fold correction factor to estimate RSV incidence among hospitalized older adults. These findings are consistent with pairwise comparisons from a recent systematic literature review and meta-analysis where, when combining results for all adults and across all treatment settings, adding one additional specimen type to nasal/NPS RT-PCR increased RSV detection by 52% for sputum and 44% for paired serology [13]. In this review, no studies were found that looked directly at the relationship between NPS/nasal swab and saliva, but a 28% increase was reported with addition of an oropharyngeal swab RT-PCR.

The current study found greater utility of paired serology than the meta-analysis, possibly because the latter used studies that included a mix of serology methods, with different antigens in each. The non-vaccine antigen RSV assay used here contains recombinant antigens and peptides which help in reducing assay background false positivity typically seen in assays that use viral lysates or partially purified proteins [20].

Multi-specimen testing was particularly useful for identifying RSV in persons hospitalized with cardiac conditions. A growing body of literature has reported a relationship between respiratory infections and cardiac disease and more specifically, with RSV and CHF exacerbations, arrythmia, and myocardial infarctions/ischemic disease [21–26]. Further, time-series studies have documented RSV infection association with cardiac hospitalizations and mortality. For example, recent time-series studies from Germany and Spain have shown a 1.7-to 2-fold increase in the estimated RSV-attributable hospitalization burden among older adults when including cardiac and respiratory hospitalizations rather than just respiratory [27, 28]. The finding of the higher increased detection rates in participants with cardiac diagnoses was unexpected and may reflect a different infection timing relative to disease or to presentation for care. RSV infection prompting such events may be more distant from hospital admission, causing patients to have less virus present in upper airways or less nasal secretions available to capture with an NPS at the time of admission. Although we did not collect medication histories, potential less aggressive nasal sampling of participants on anticoagulants may also be contributing.

While serology and sputum had the highest sensitivities of specimen types, completeness of collection was an issue. Many participants were unable to produce sputum, and serology is less useful in the clinical setting because convalescent sera collection is required, such that results arrive too late to influence patient care. Saliva/saline mouthwash is a more practical additional specimen; among the specimen types collected, saliva identified the highest number of RSV positive specimens since nearly all participants provided specimens and saliva had higher sensitivity (63% compared to 49% for NPS). However, current clinical practice and SOC testing for respiratory illness among adults hospitalized for ARI do not routinely include saliva sampling despite recent studies demonstrating its high sensitivity and specificity when compared to NPS/aspirates for RSV [29], influenza [30], and similar sensitivity to NPS for SARS-CoV-2 detection [31], and its recent validation for identification of RSV and other viral pathogens [32].

In participants with paired swabs available, we documented higher rates of RSV detection from earlier swabs compared to later swabs. This finding is likely due to the former being collected on average one day earlier. Viral load is known to fall quickly in respiratory secretions in adults [12]. Thus, rapid specimen collection in adult patients presenting with ARI is critical and use of salvaged SOC swabs or sputa for burden studies wherever possible should be included on study protocols. Studies depending on research specimen collection only could see a reduction of burden of nearly one-third (30%) with only a day delay in specimen collection from presentation.

Our study had several limitations. Not all subjects had all four specimen types collected. Half of the cohort did not complete the final study visit for convalescent sera collection, highlighting the difficulty of using this specimen type. Study assay performance was not defined for every specimen type thus the rate of false positive detection may not be known, but specificity for all test types is known to be high (>99%) [13]. The study was conducted during the latter part of the COVID-19 pandemic, which may have altered the epidemiology of RSV infection, although there is no reason to suspect that it would change the relative sensitivity of different specimen types for diagnosis. Lastly, sites were limited to two geographic areas in North America, and specimen collection practices may differ globally, although pairwise results from around the world have shown similar trends in a recent global systematic review [13].

Despite different participant profiles in US and Canadian sites, both settings demonstrated substantially higher RSV detection when specimens were obtained beyond NPS, with all subpopulations benefiting from the collection of additional specimens. Our data confirm that recent meta-analyses application of a correction factor to RSV disease burden estimates based on NPS alone can contribute to more accurate population-based estimates. Corrected rates provide more accurate information for policy makers and public health officials when assessing RSV vaccine impact and planning for future RSV antiviral use when such therapeutics have been licensed. Research studies should consider collection of salvaged respiratory specimens, which are collected earlier in the course of illness, to reduce missed infection wherever feasible. Additionally, future studies may be conducted in the setting of RSV vaccine use, which may result in breakthrough cases having lower viral load in respiratory secretions[20], thereby increasing the utility of collecting additional sample types. Finally, given the sensitivity and relative ease of collection, consideration should be given to advancing routine use of saliva for RSV testing, including its utility among certain patient subgroups such as those with cardiac diagnoses.

## Data Availability

All data produced in the present work are contained in the manuscript

## FOOTNOTES

## CONFERENCE ABSTRACT

The results of this study were presented at RSVVW’24, 8^th^ ReSViNET Conference, 13-16 February, 2024 at Mumbai, Maharashtra, India.

## DATA AVAILABILITY

The datasets analyzed during the current study are not publicly available.

## Supplementary Tables

**Supplementary Table 1.**
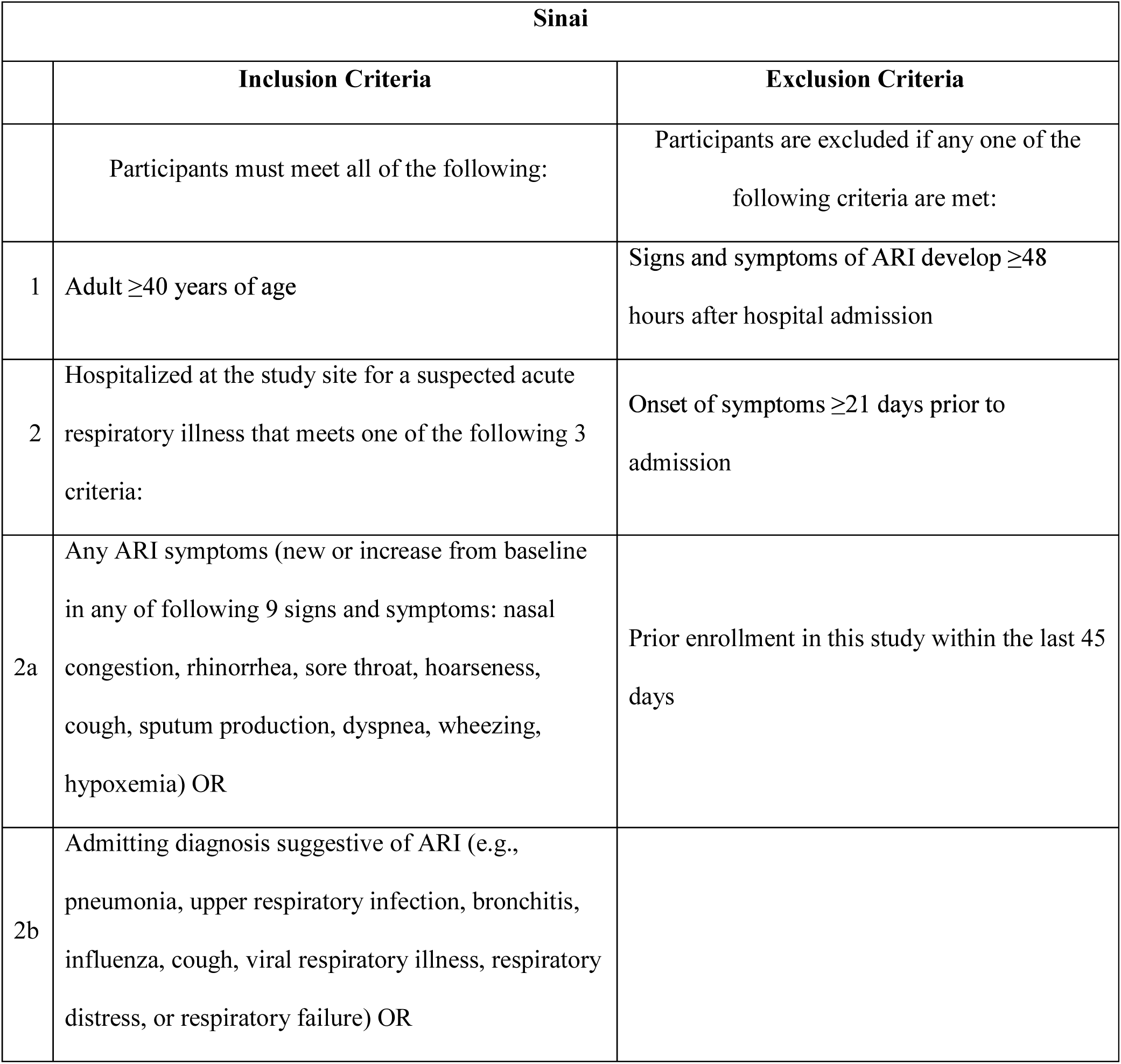
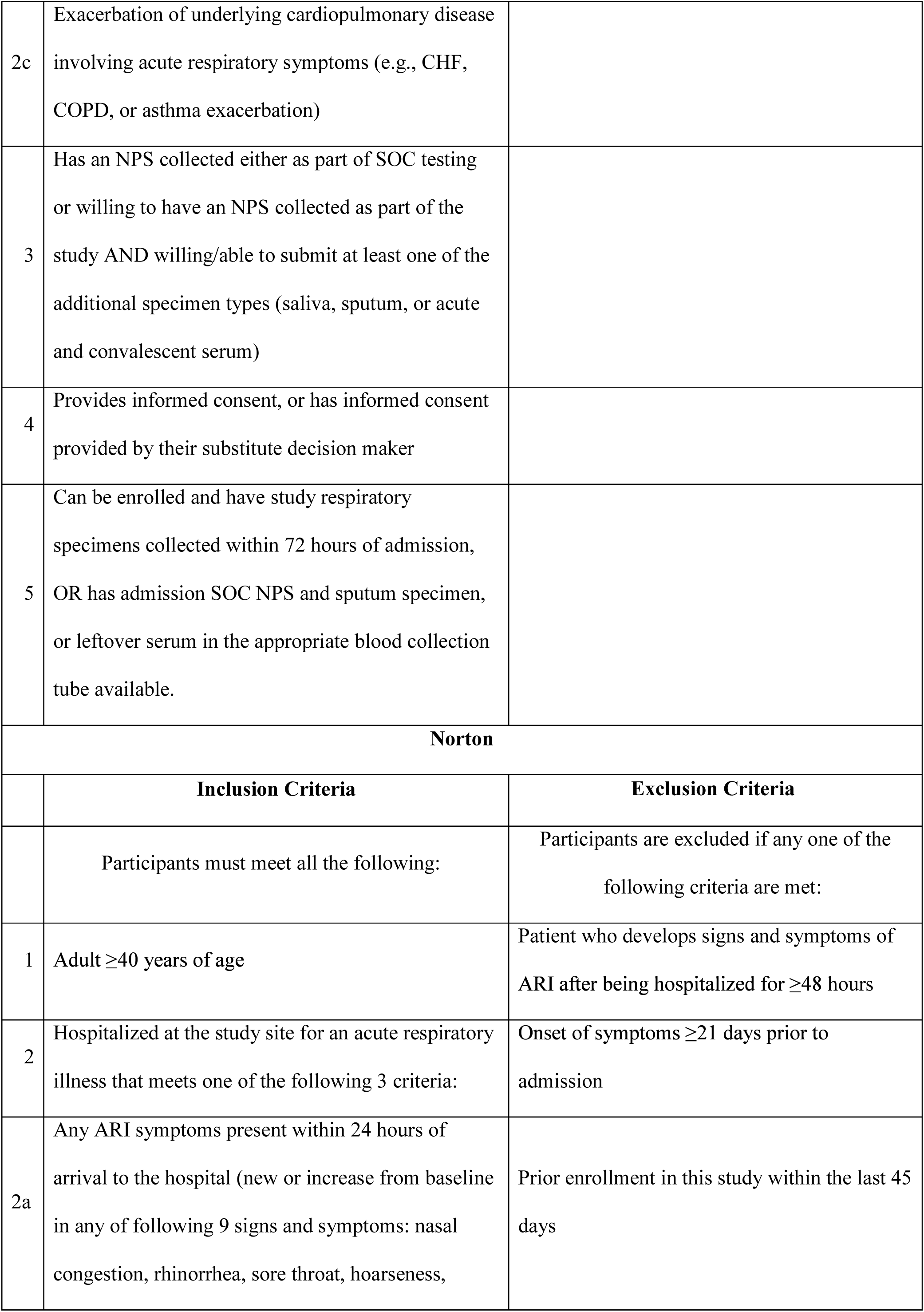
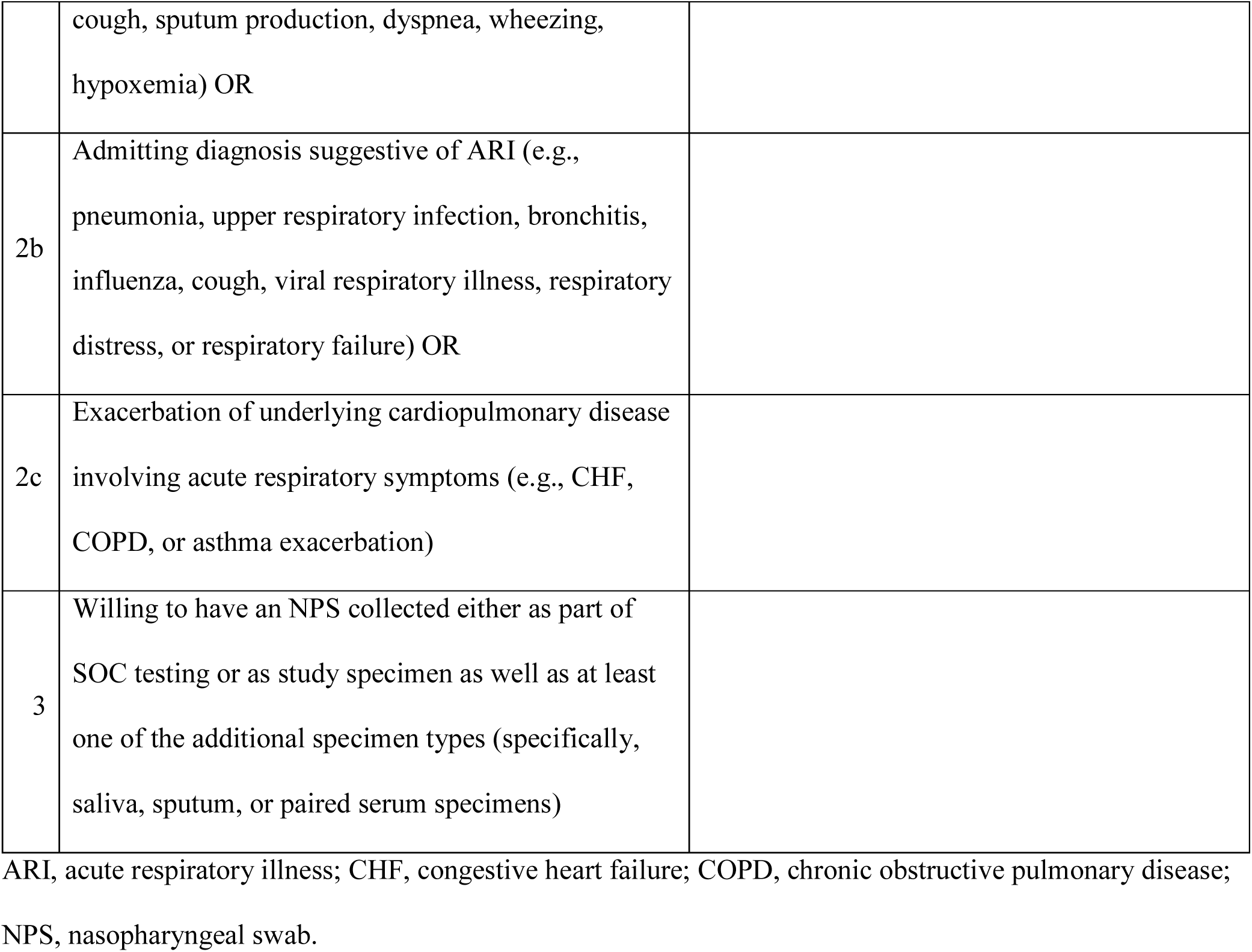
Inclusion criteria and exclusion criteria.

**Supplementary Table 2:**
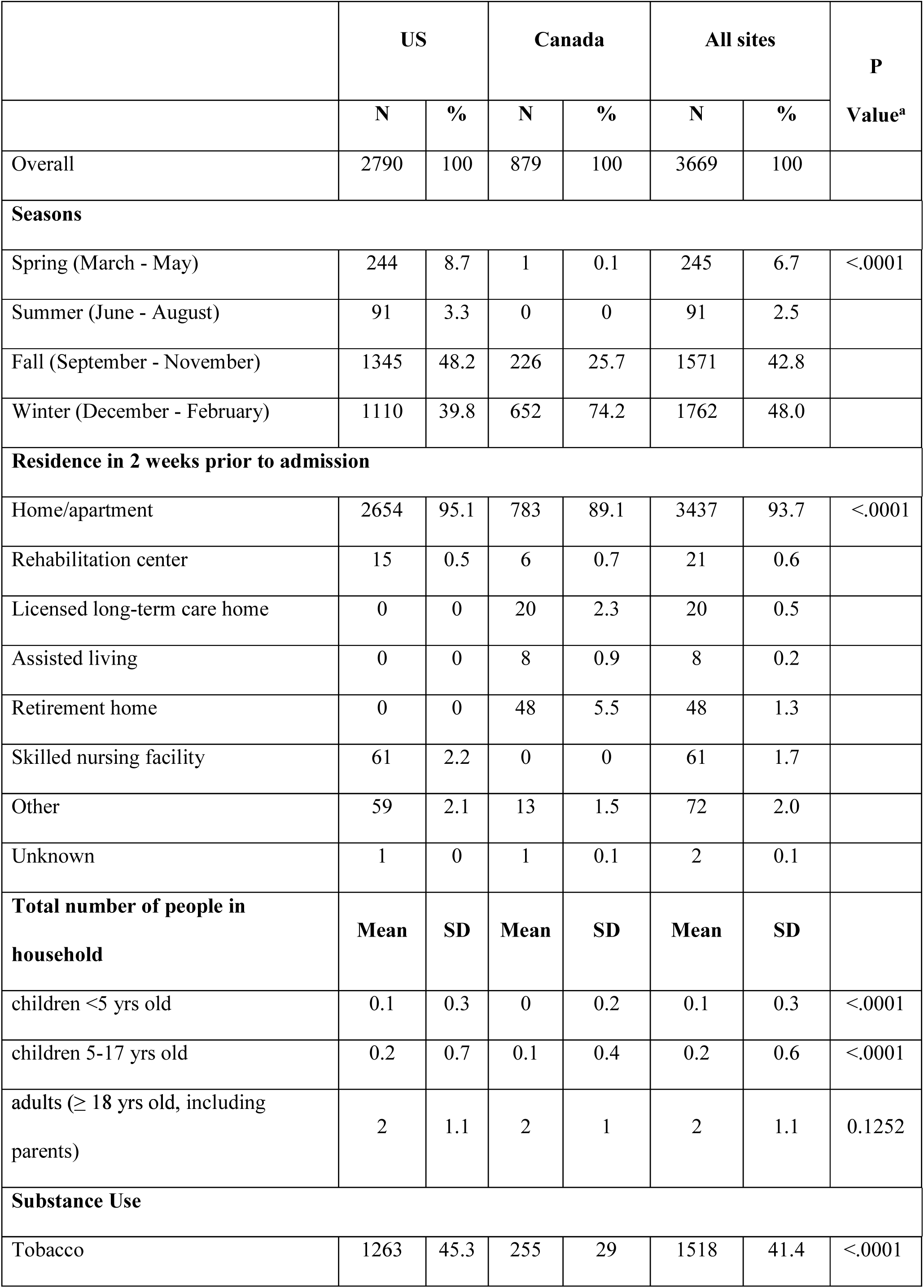
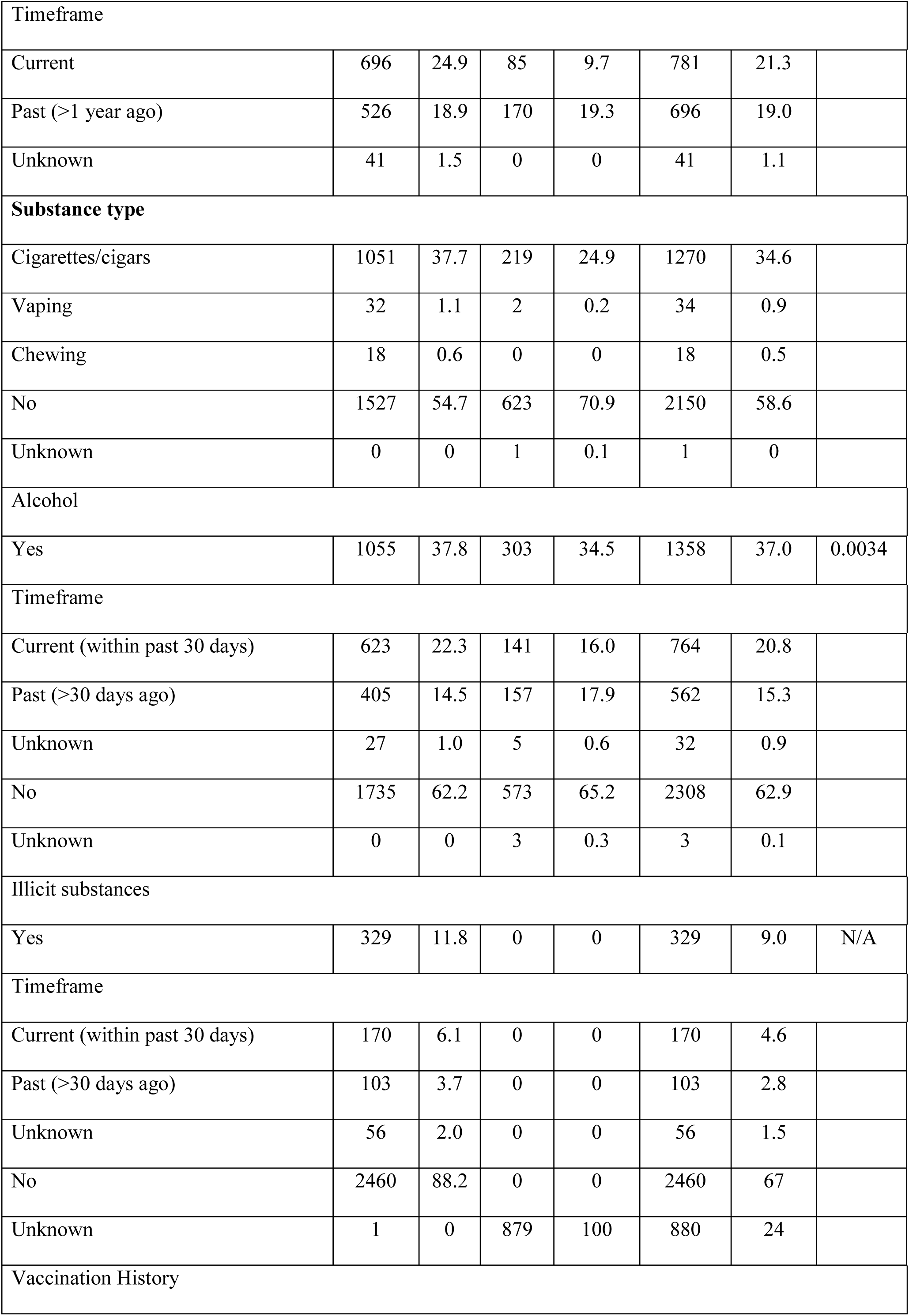
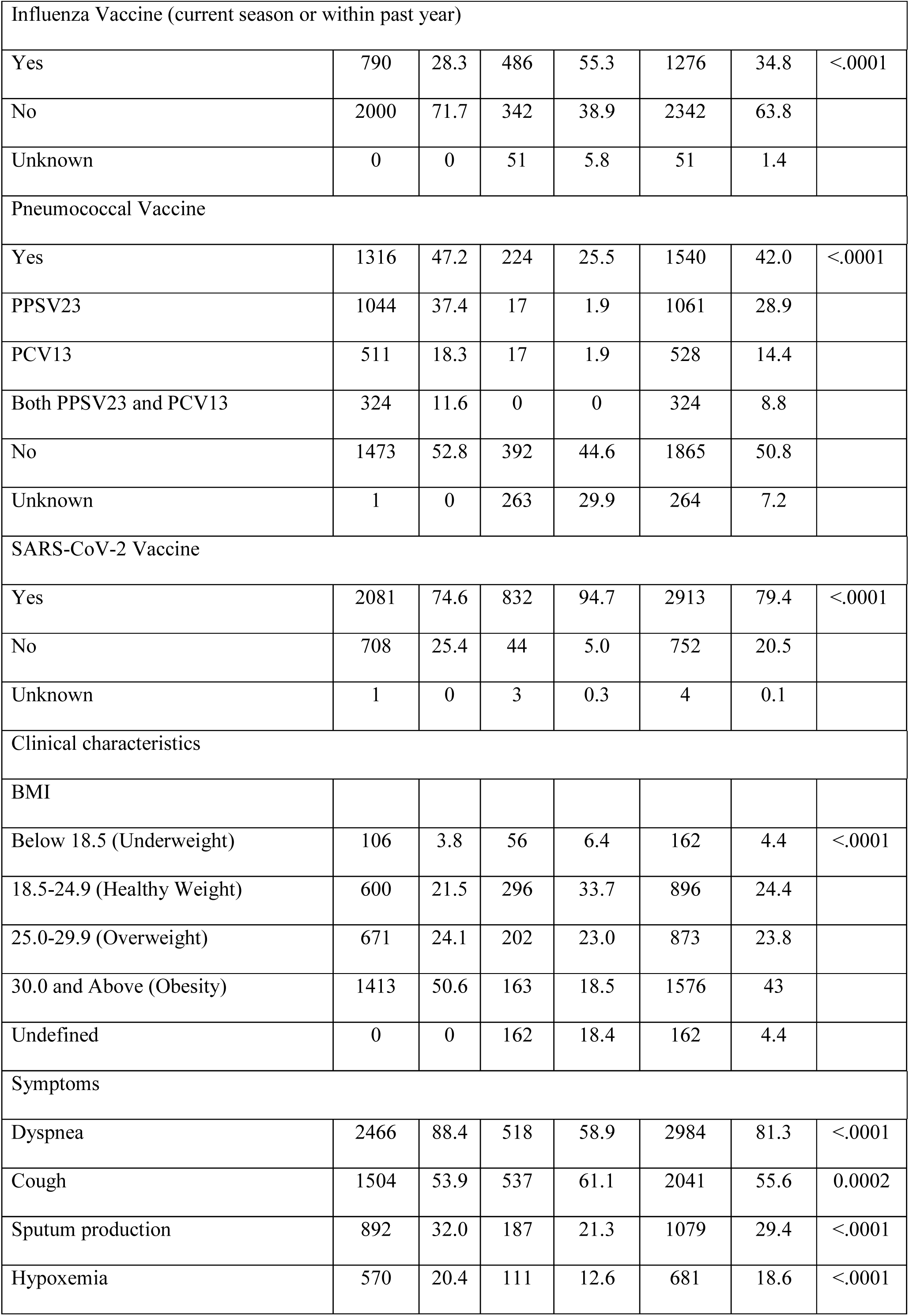
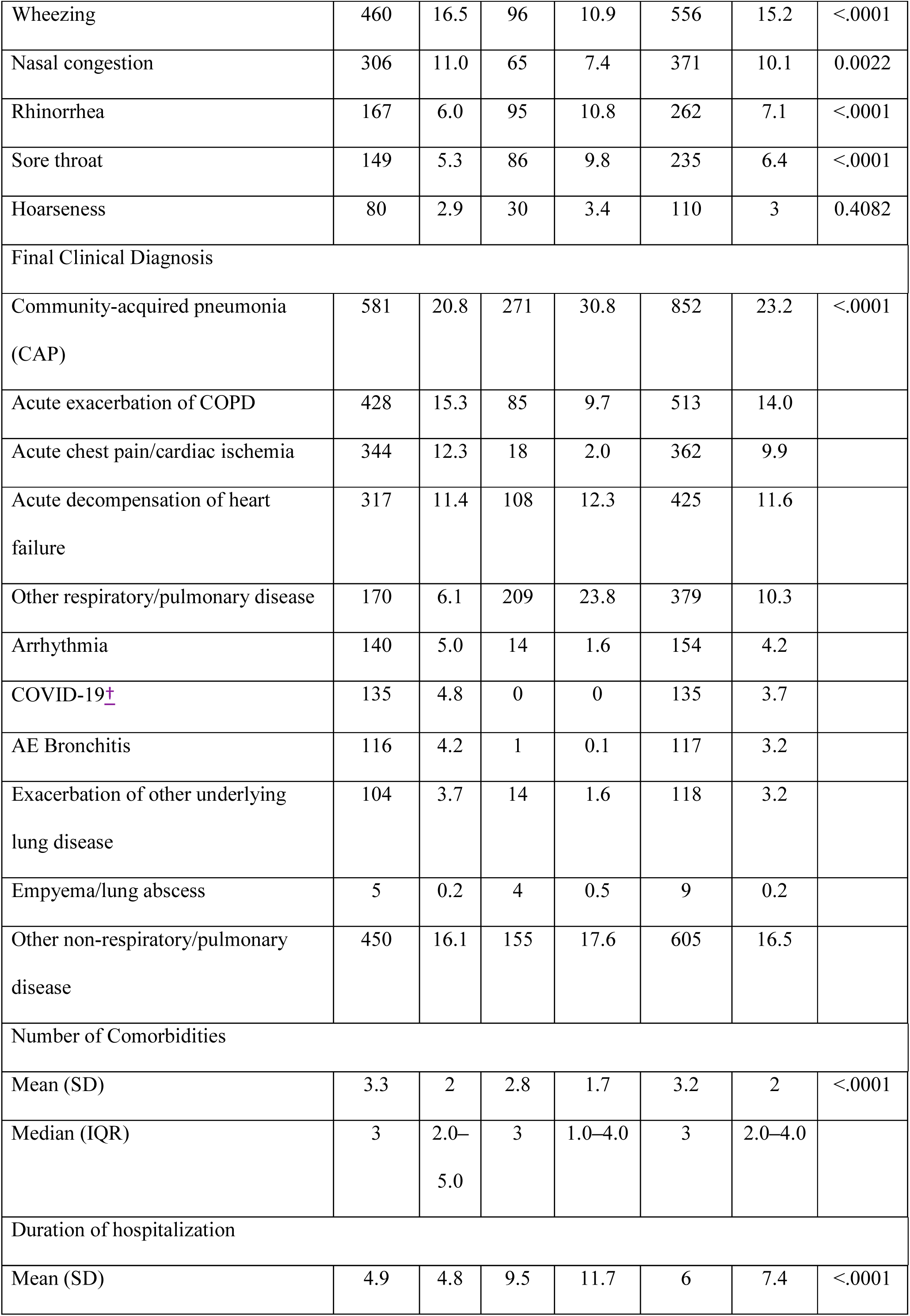
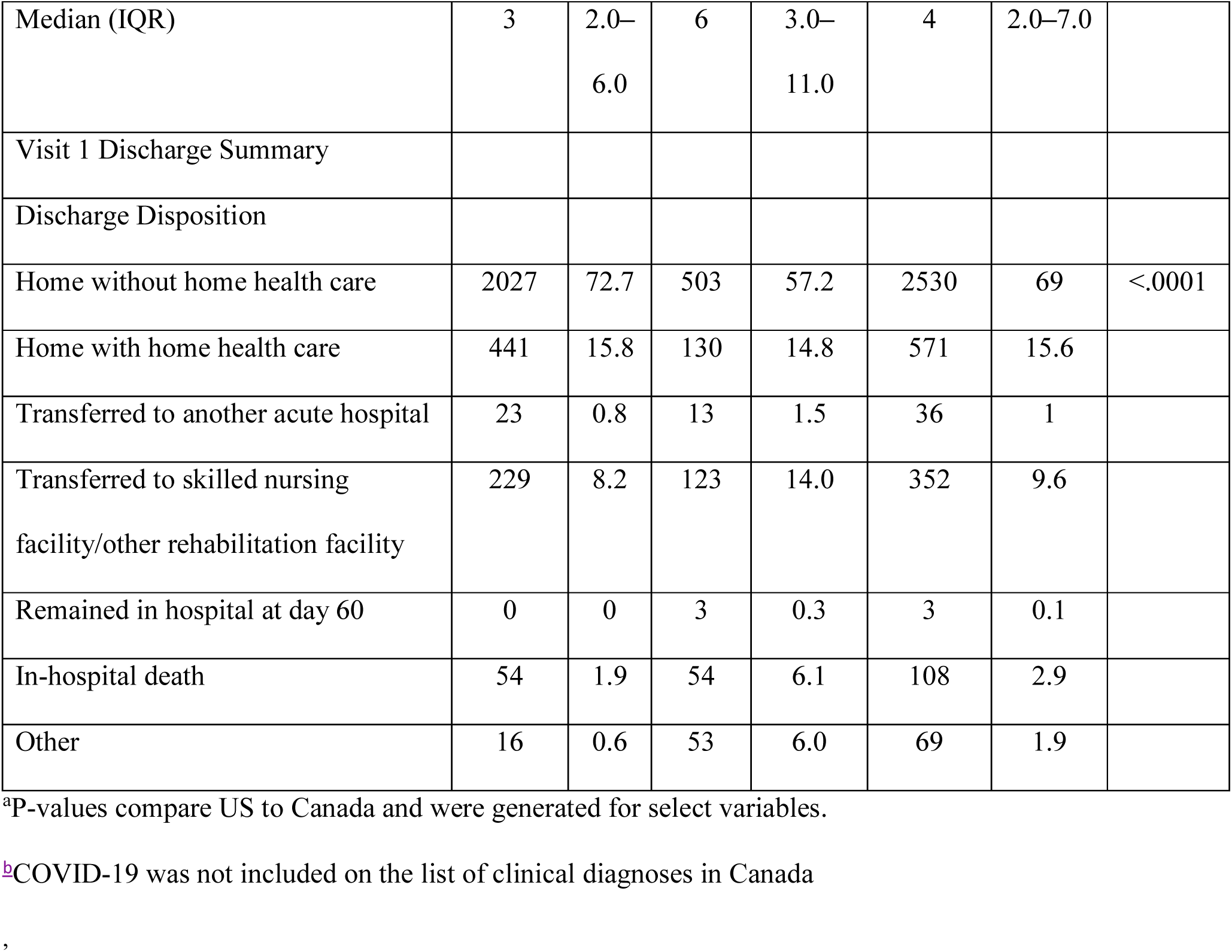
Additional characteristics of study participants, overall and by site.

**Supplementary Table 3.**
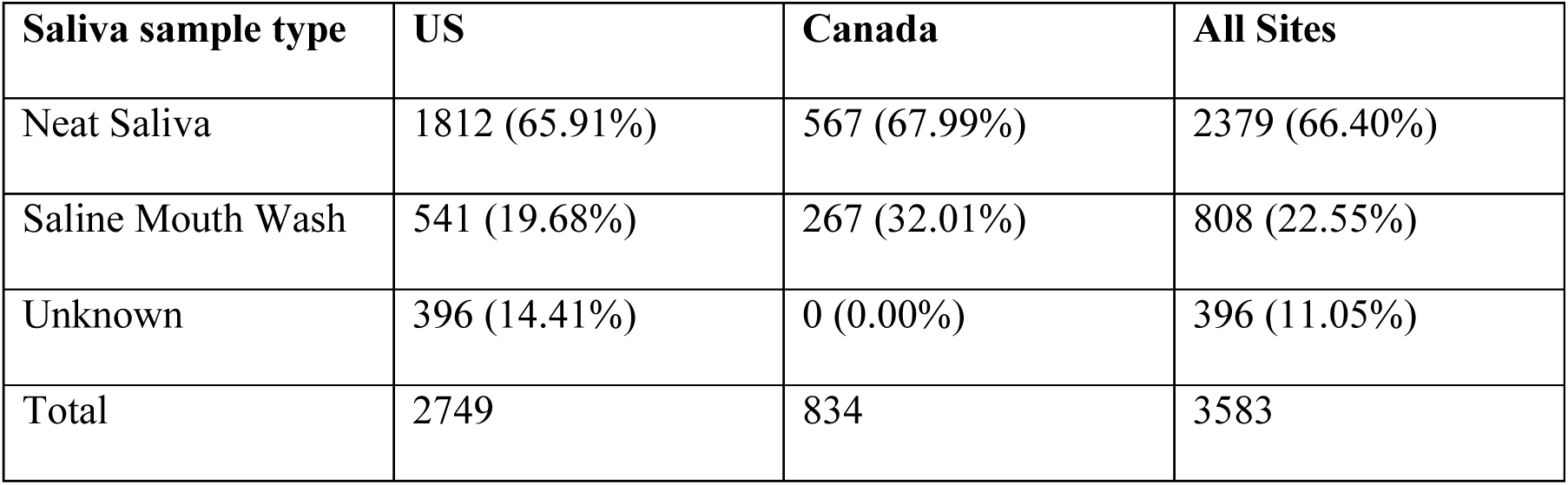
Saliva specimens by collection method.

**Supplementary Table 4:**
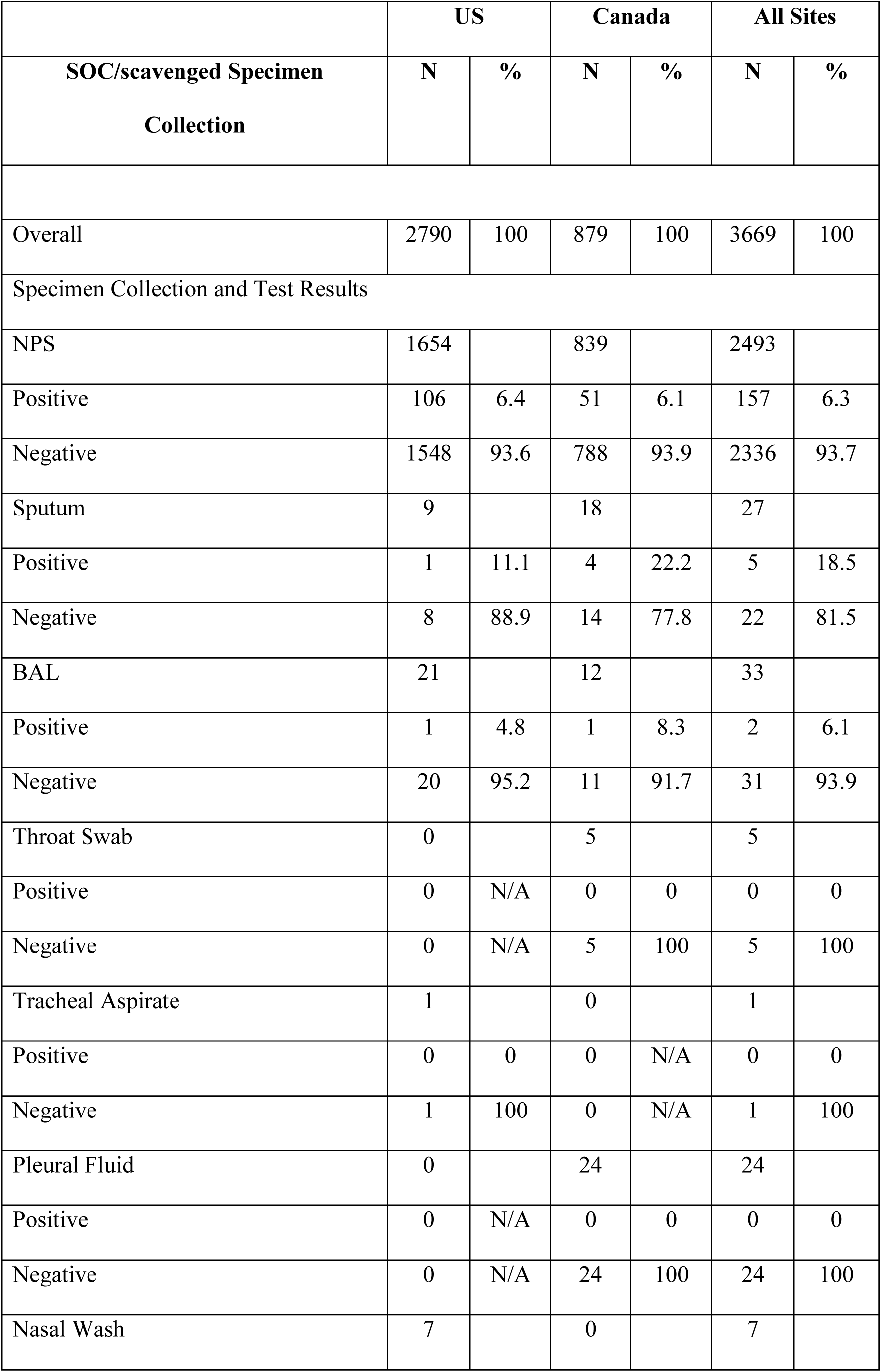
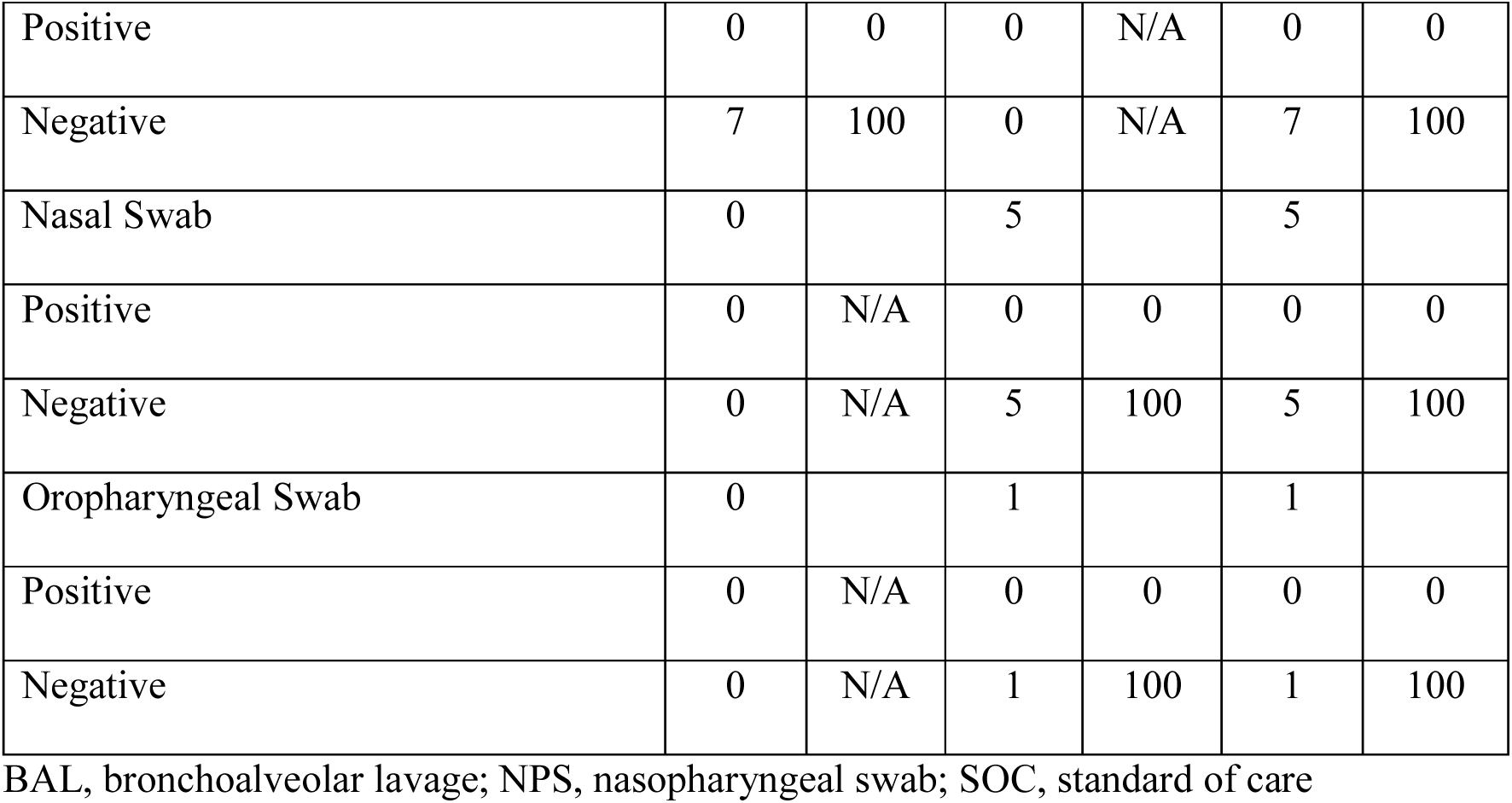
Specimen collection and RSV test results for standard of care or scavenged specimen testing.

**Supplementary Table 5:**
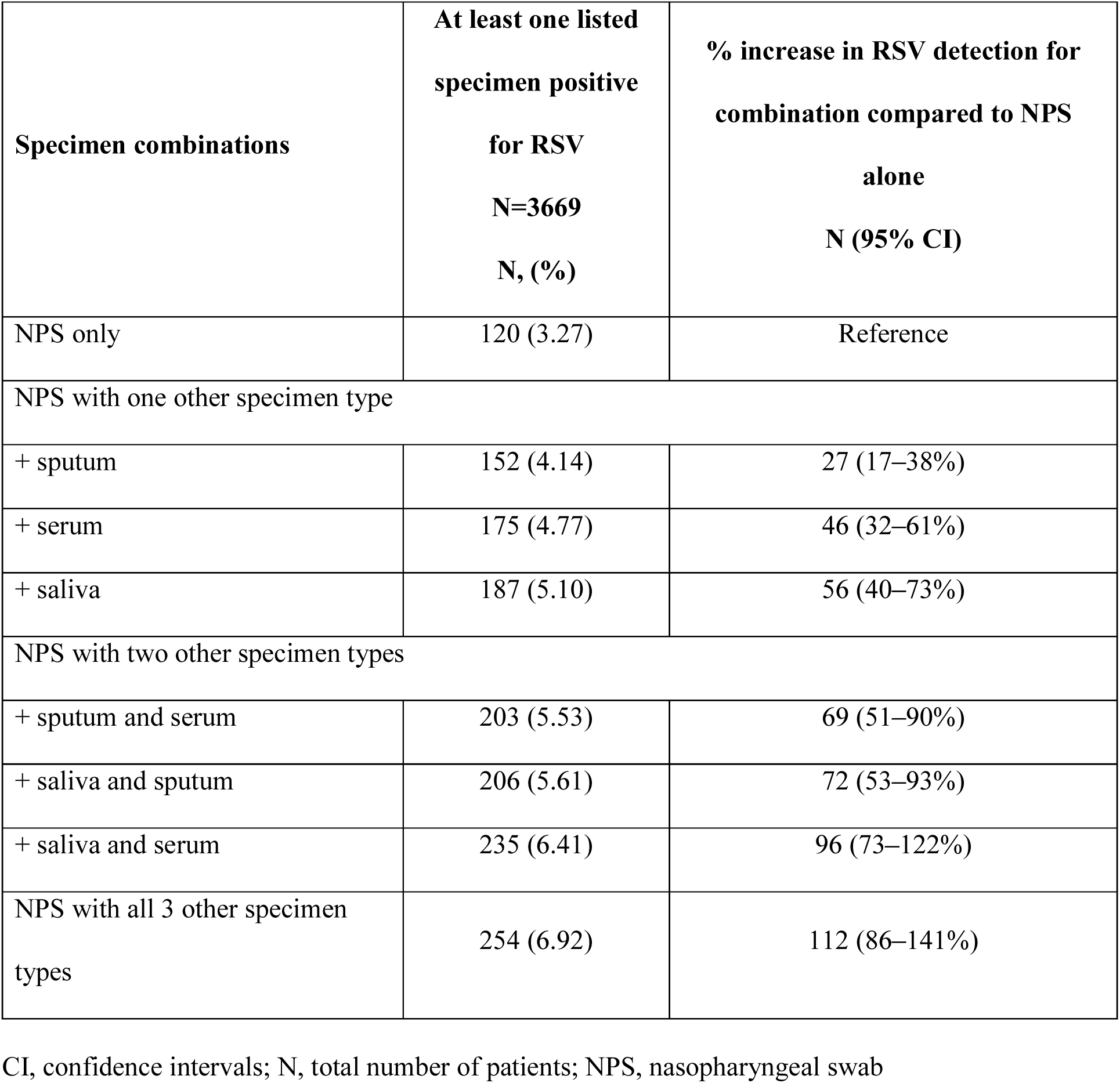
Increase in RSV detection associated with using additional specimen type results beyond NP swab for all participants in the study (N=3669)

**Supplementary Table 6:**
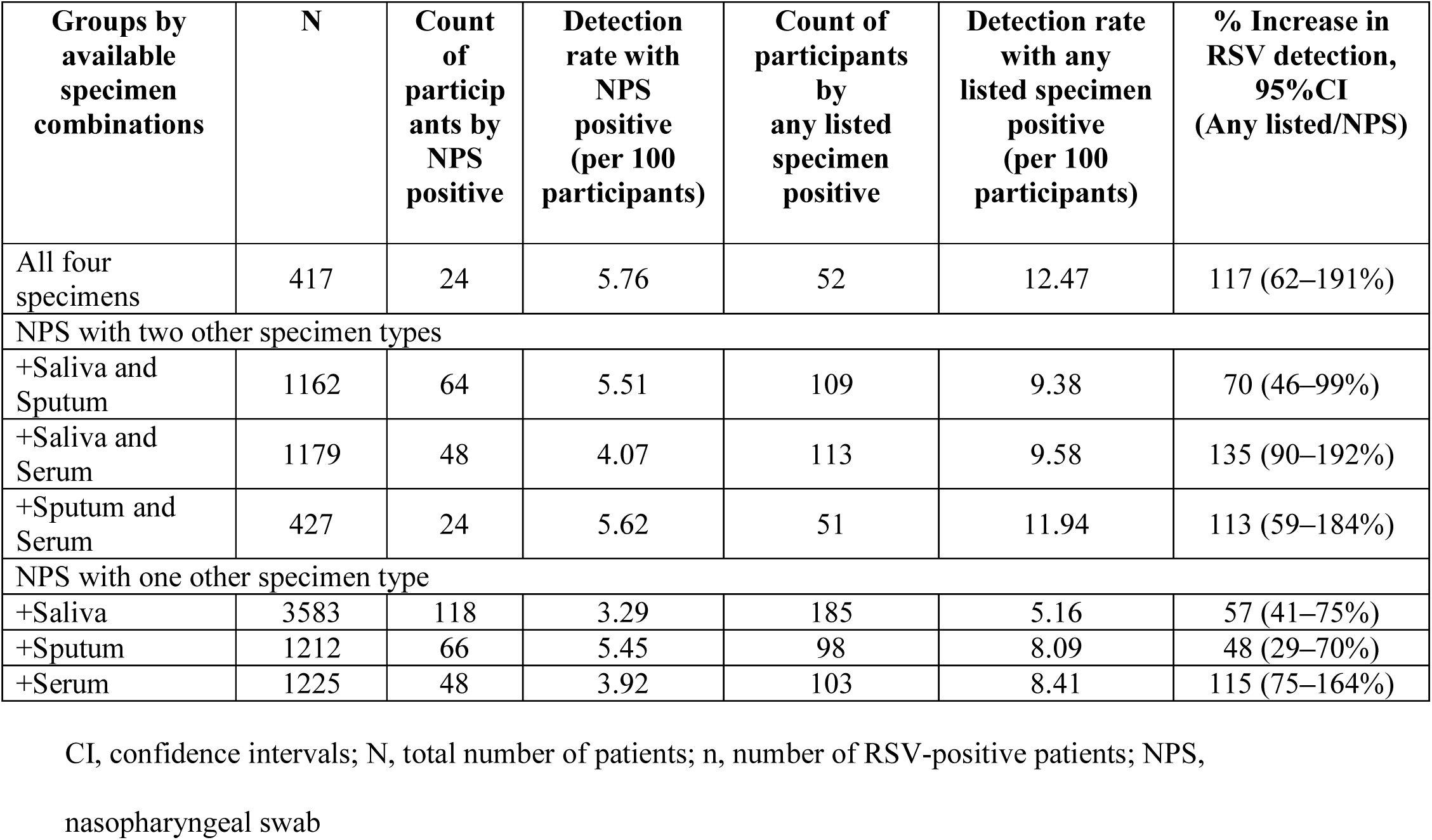
Increase in RSV detection associated with using additional specimen type results beyond NP swab for different populations with specific specimen results available (N variable per specimen type combination)

**Supplementary Table 7:**
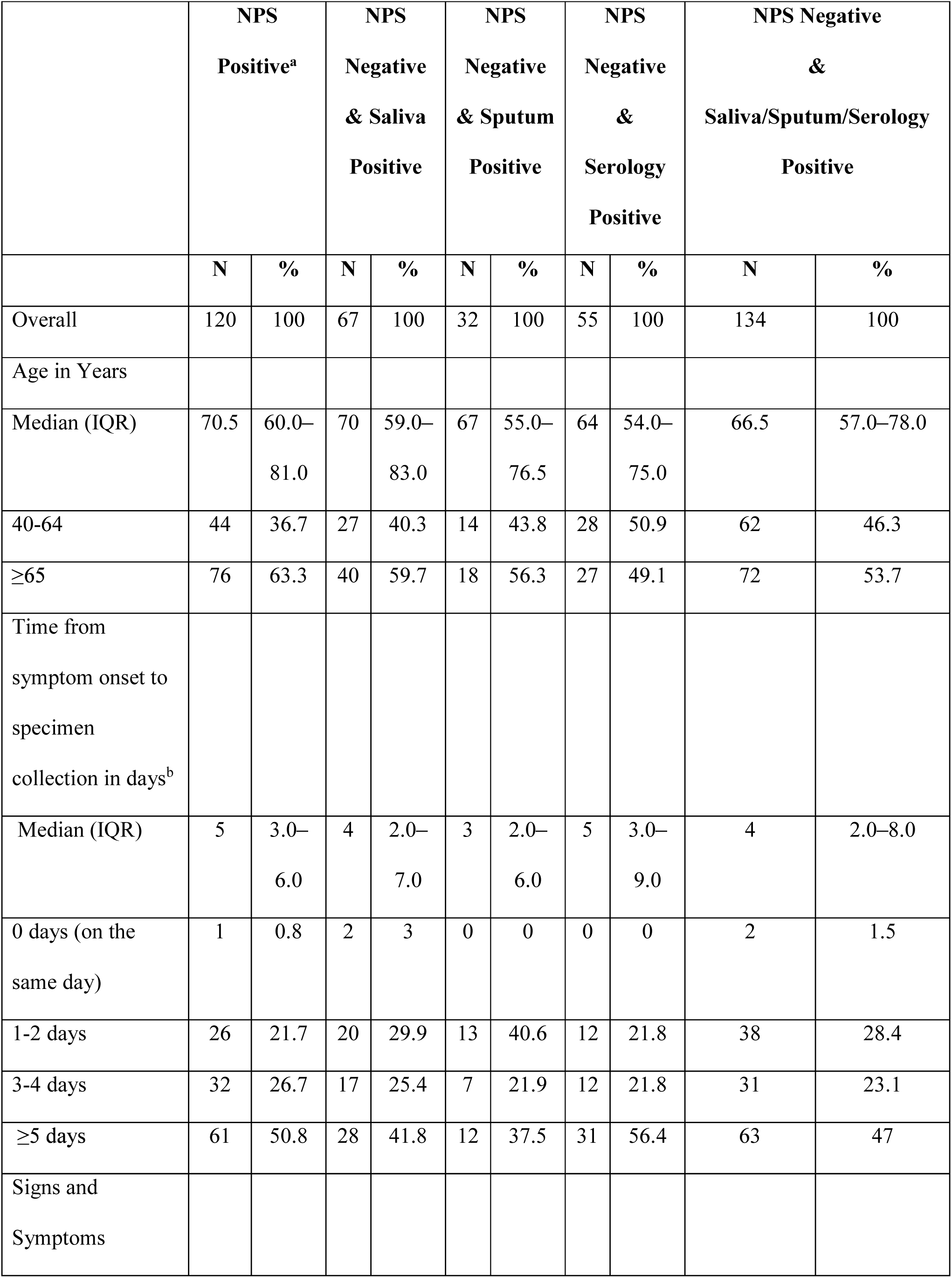
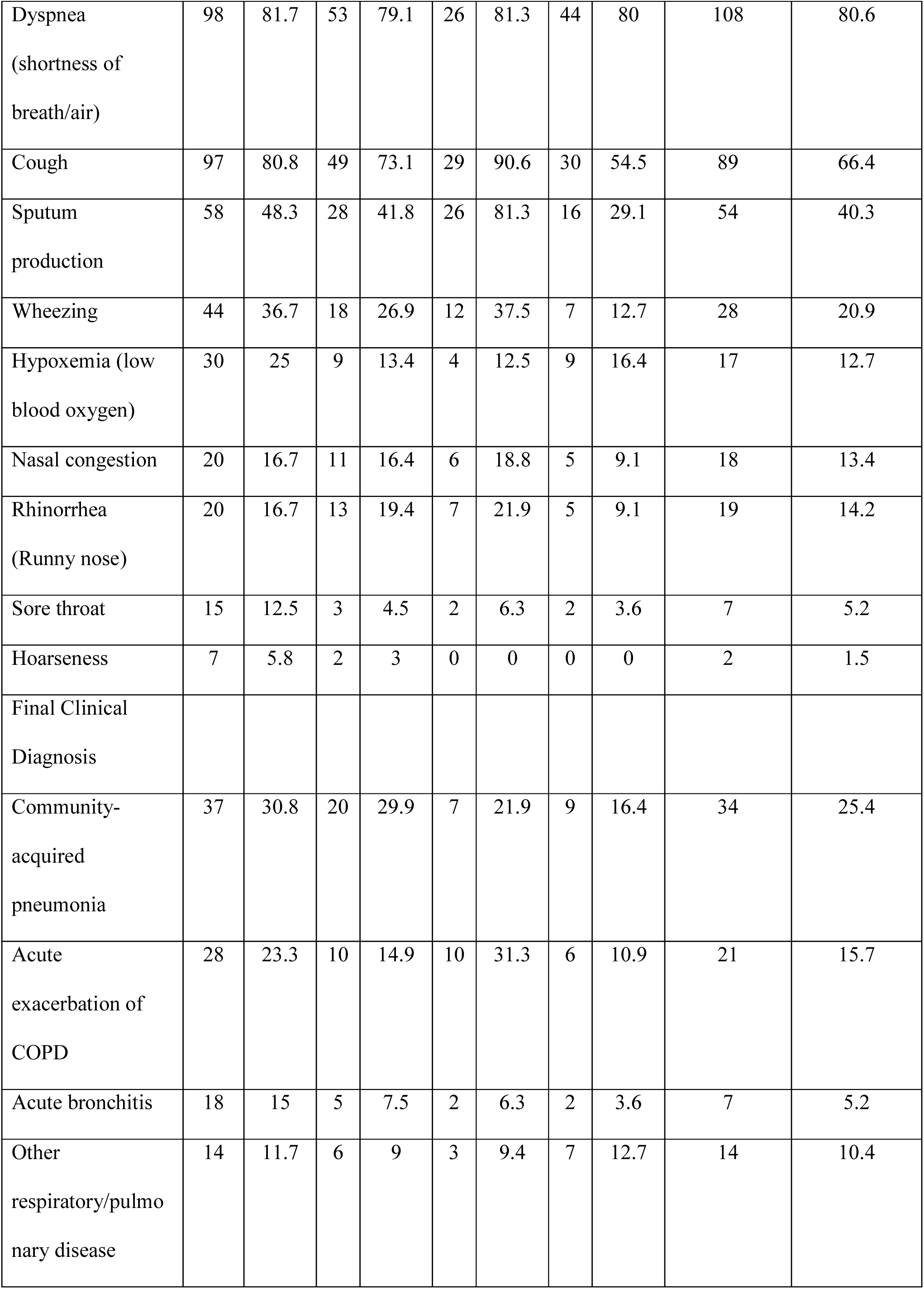
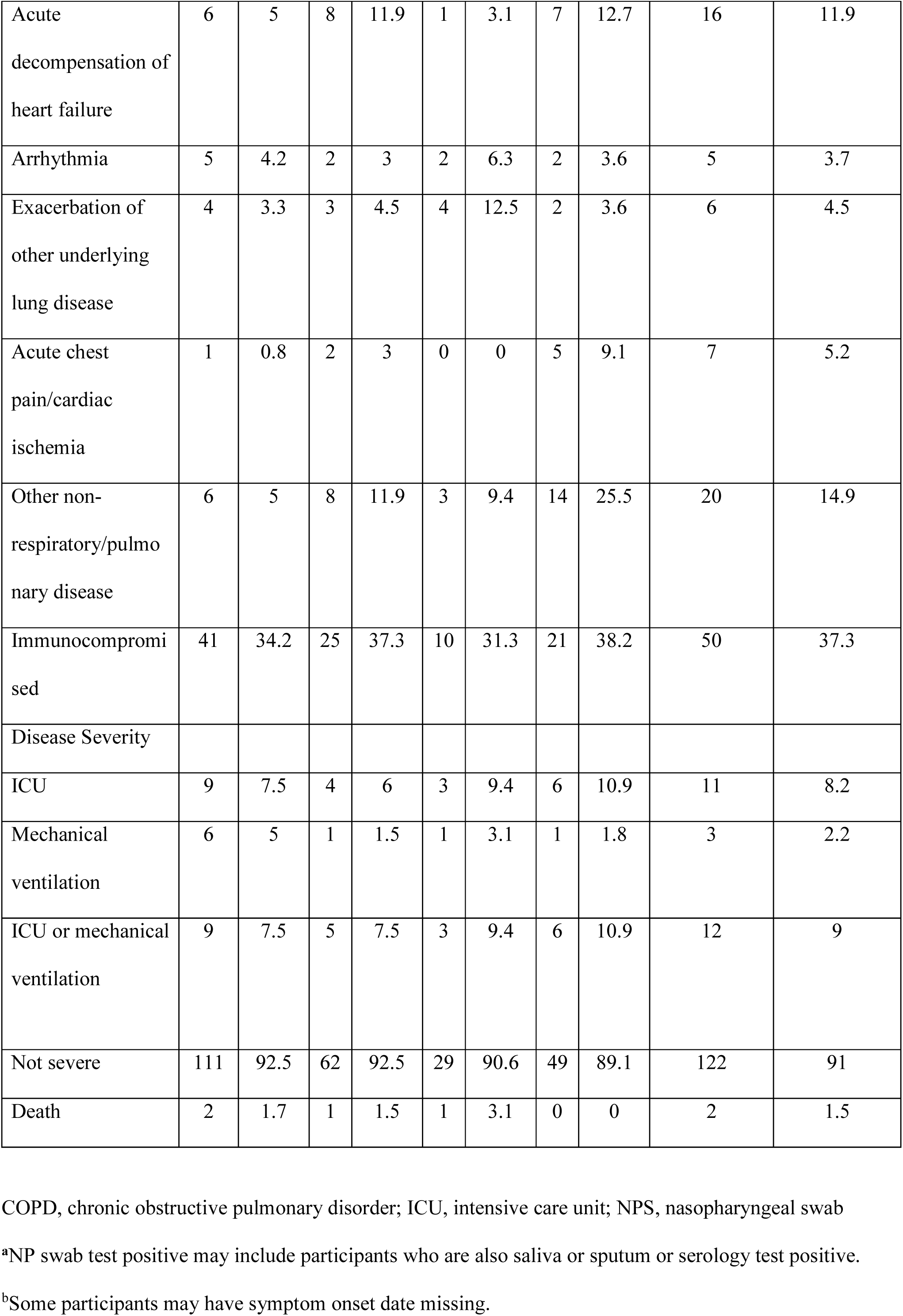
Characteristics of RSV participants identified by NPS compared with RSV participants identified by other specimen types only.

