## supplemental files for "Detection of RSV using nasopharyngeal swabs alone underestimates RSV-related hospitalization incidence in adults: the Multispecimen study’s Final Analysis"

**Supplementary Tables**

**Supplementary Table 1. Inclusion criteria and exclusion criteria**

| Sinai | | |
|---|---|---|
| | **Inclusion Criteria** | **Exclusion Criteria** |
| | Participants must meet all of the following: | Participants are excluded if any one of the following criteria are met: |
| 1 | Adult ≥40 years of age | Signs and symptoms of ARI develop ≥48 hours after hospital admission |
| 2 | Hospitalized at the study site for a suspected acute respiratory illness that meets one of the following 3 criteria: | Onset of symptoms ≥21 days prior to admission |
| 2a | Any ARI symptoms (new or increase from baseline in any of following 9 signs and symptoms: nasal congestion, rhinorrhea, sore throat, hoarseness, cough, sputum production, dyspnea, wheezing, hypoxemia) OR | Prior enrollment in this study within the last 45 days |
| 2b | Admitting diagnosis suggestive of ARI (e.g., pneumonia, upper respiratory infection, bronchitis, influenza, cough, viral respiratory illness, respiratory distress, or respiratory failure) OR | |

| | | |
|---|---|---|
| 2c | Exacerbation of underlying cardiopulmonary disease involving acute respiratory symptoms (e.g., CHF, COPD, or asthma exacerbation) | |
| 3 | Has an NPS collected either as part of SOC testing or willing to have an NPS collected as part of the study AND willing/able to submit at least one of the additional specimen types (saliva, sputum, or acute and convalescent serum) | |
| 4 | Provides informed consent, or has informed consent provided by their substitute decision maker | |
| 5 | Can be enrolled and have study respiratory specimens collected within 72 hours of admission, OR has admission SOC NPS and sputum specimen, or leftover serum in the appropriate blood collection tube available. | |
| | **Norton** | |
| | **Inclusion Criteria** | **Exclusion Criteria** |
| | Participants must meet all the following: | Participants are excluded if any one of the following criteria are met: |
| 1 | Adult ≥40 years of age | Patient who develops signs and symptoms of ARI after being hospitalized for ≥48 hours |
| 2 | Hospitalized at the study site for an acute respiratory illness that meets one of the following 3 criteria: | Onset of symptoms ≥21 days prior to admission |
| 2a | Any ARI symptoms present within 24 hours of arrival to the hospital (new or increase from baseline in any of following 9 signs and symptoms: nasal congestion, rhinorrhea, sore throat, hoarseness, | Prior enrollment in this study within the last 45 days |

| | |
|---|---|
| | cough, sputum production, dyspnea, wheezing, hypoxemia) OR |
| 2b | Admitting diagnosis suggestive of ARI (e.g., pneumonia, upper respiratory infection, bronchitis, influenza, cough, viral respiratory illness, respiratory distress, or respiratory failure) OR |
| 2c | Exacerbation of underlying cardiopulmonary disease involving acute respiratory symptoms (e.g., CHF, COPD, or asthma exacerbation) |
| 3 | Willing to have an NPS collected either as part of SOC testing or as study specimen as well as at least one of the additional specimen types (specifically, saliva, sputum, or paired serum specimens) |

ARI, acute respiratory illness; CHF, congestive heart failure; COPD, chronic obstructive pulmonary disease; NPS, nasopharyngeal swab.

**Supplementary Table 2: Additional characteristics of study participants, overall and by site**

| | US | | Canada | | All sites | | P Value[a] |
|---|---|---|---|---|---|---|---|
| | N | % | N | % | N | % | |
| Overall | 2790 | 100 | 879 | 100 | 3669 | 100 | |
| **Seasons** | | | | | | | |
| Spring (March - May) | 244 | 8.7 | 1 | 0.1 | 245 | 6.7 | <.0001 |
| Summer (June - August) | 91 | 3.3 | 0 | 0 | 91 | 2.5 | |
| Fall (September - November) | 1345 | 48.2 | 226 | 25.7 | 1571 | 42.8 | |
| Winter (December - February) | 1110 | 39.8 | 652 | 74.2 | 1762 | 48.0 | |
| **Residence in 2 weeks prior to admission** | | | | | | | |
| Home/apartment | 2654 | 95.1 | 783 | 89.1 | 3437 | 93.7 | <.0001 |
| Rehabilitation center | 15 | 0.5 | 6 | 0.7 | 21 | 0.6 | |
| Licensed long-term care home | 0 | 0 | 20 | 2.3 | 20 | 0.5 | |
| Assisted living | 0 | 0 | 8 | 0.9 | 8 | 0.2 | |
| Retirement home | 0 | 0 | 48 | 5.5 | 48 | 1.3 | |
| Skilled nursing facility | 61 | 2.2 | 0 | 0 | 61 | 1.7 | |
| Other | 59 | 2.1 | 13 | 1.5 | 72 | 2.0 | |
| Unknown | 1 | 0 | 1 | 0.1 | 2 | 0.1 | |
| **Total number of people in household** | **Mean** | **SD** | **Mean** | **SD** | **Mean** | **SD** | |
| children <5 yrs old | 0.1 | 0.3 | 0 | 0.2 | 0.1 | 0.3 | <.0001 |
| children 5-17 yrs old | 0.2 | 0.7 | 0.1 | 0.4 | 0.2 | 0.6 | <.0001 |
| adults (≥ 18 yrs old, including parents) | 2 | 1.1 | 2 | 1 | 2 | 1.1 | 0.1252 |
| **Substance Use** | | | | | | | |
| Tobacco | 1263 | 45.3 | 255 | 29 | 1518 | 41.4 | <.0001 |

| | | | | | | | |
|---|---|---|---|---|---|---|---|
| Timeframe | | | | | | | |
| Current | 696 | 24.9 | 85 | 9.7 | 781 | 21.3 | |
| Past (>1 year ago) | 526 | 18.9 | 170 | 19.3 | 696 | 19.0 | |
| Unknown | 41 | 1.5 | 0 | 0 | 41 | 1.1 | |
| **Substance type** | | | | | | | |
| Cigarettes/cigars | 1051 | 37.7 | 219 | 24.9 | 1270 | 34.6 | |
| Vaping | 32 | 1.1 | 2 | 0.2 | 34 | 0.9 | |
| Chewing | 18 | 0.6 | 0 | 0 | 18 | 0.5 | |
| No | 1527 | 54.7 | 623 | 70.9 | 2150 | 58.6 | |
| Unknown | 0 | 0 | 1 | 0.1 | 1 | 0 | |
| Alcohol | | | | | | | |
| Yes | 1055 | 37.8 | 303 | 34.5 | 1358 | 37.0 | 0.0034 |
| Timeframe | | | | | | | |
| Current (within past 30 days) | 623 | 22.3 | 141 | 16.0 | 764 | 20.8 | |
| Past (>30 days ago) | 405 | 14.5 | 157 | 17.9 | 562 | 15.3 | |
| Unknown | 27 | 1.0 | 5 | 0.6 | 32 | 0.9 | |
| No | 1735 | 62.2 | 573 | 65.2 | 2308 | 62.9 | |
| Unknown | 0 | 0 | 3 | 0.3 | 3 | 0.1 | |
| Illicit substances | | | | | | | |
| Yes | 329 | 11.8 | 0 | 0 | 329 | 9.0 | N/A |
| Timeframe | | | | | | | |
| Current (within past 30 days) | 170 | 6.1 | 0 | 0 | 170 | 4.6 | |
| Past (>30 days ago) | 103 | 3.7 | 0 | 0 | 103 | 2.8 | |
| Unknown | 56 | 2.0 | 0 | 0 | 56 | 1.5 | |
| No | 2460 | 88.2 | 0 | 0 | 2460 | 67 | |
| Unknown | 1 | 0 | 879 | 100 | 880 | 24 | |
| Vaccination History | | | | | | | |

| | | | | | | | |
|---|---|---|---|---|---|---|---|
| Influenza Vaccine (current season or within past year) | | | | | | | |
| Yes | 790 | 28.3 | 486 | 55.3 | 1276 | 34.8 | <.0001 |
| No | 2000 | 71.7 | 342 | 38.9 | 2342 | 63.8 | |
| Unknown | 0 | 0 | 51 | 5.8 | 51 | 1.4 | |
| Pneumococcal Vaccine | | | | | | | |
| Yes | 1316 | 47.2 | 224 | 25.5 | 1540 | 42.0 | <.0001 |
| PPSV23 | 1044 | 37.4 | 17 | 1.9 | 1061 | 28.9 | |
| PCV13 | 511 | 18.3 | 17 | 1.9 | 528 | 14.4 | |
| Both PPSV23 and PCV13 | 324 | 11.6 | 0 | 0 | 324 | 8.8 | |
| No | 1473 | 52.8 | 392 | 44.6 | 1865 | 50.8 | |
| Unknown | 1 | 0 | 263 | 29.9 | 264 | 7.2 | |
| SARS-CoV-2 Vaccine | | | | | | | |
| Yes | 2081 | 74.6 | 832 | 94.7 | 2913 | 79.4 | <.0001 |
| No | 708 | 25.4 | 44 | 5.0 | 752 | 20.5 | |
| Unknown | 1 | 0 | 3 | 0.3 | 4 | 0.1 | |
| Clinical characteristics | | | | | | | |
| BMI | | | | | | | |
| Below 18.5 (Underweight) | 106 | 3.8 | 56 | 6.4 | 162 | 4.4 | <.0001 |
| 18.5-24.9 (Healthy Weight) | 600 | 21.5 | 296 | 33.7 | 896 | 24.4 | |
| 25.0-29.9 (Overweight) | 671 | 24.1 | 202 | 23.0 | 873 | 23.8 | |
| 30.0 and Above (Obesity) | 1413 | 50.6 | 163 | 18.5 | 1576 | 43 | |
| Undefined | 0 | 0 | 162 | 18.4 | 162 | 4.4 | |
| Symptoms | | | | | | | |
| Dyspnea | 2466 | 88.4 | 518 | 58.9 | 2984 | 81.3 | <.0001 |
| Cough | 1504 | 53.9 | 537 | 61.1 | 2041 | 55.6 | 0.0002 |
| Sputum production | 892 | 32.0 | 187 | 21.3 | 1079 | 29.4 | <.0001 |
| Hypoxemia | 570 | 20.4 | 111 | 12.6 | 681 | 18.6 | <.0001 |

| | | | | | | | |
|---|---|---|---|---|---|---|---|
| Wheezing | 460 | 16.5 | 96 | 10.9 | 556 | 15.2 | <.0001 |
| Nasal congestion | 306 | 11.0 | 65 | 7.4 | 371 | 10.1 | 0.0022 |
| Rhinorrhea | 167 | 6.0 | 95 | 10.8 | 262 | 7.1 | <.0001 |
| Sore throat | 149 | 5.3 | 86 | 9.8 | 235 | 6.4 | <.0001 |
| Hoarseness | 80 | 2.9 | 30 | 3.4 | 110 | 3 | 0.4082 |
| Final Clinical Diagnosis | | | | | | | |
| Community-acquired pneumonia (CAP) | 581 | 20.8 | 271 | 30.8 | 852 | 23.2 | <.0001 |
| Acute exacerbation of COPD | 428 | 15.3 | 85 | 9.7 | 513 | 14.0 | |
| Acute chest pain/cardiac ischemia | 344 | 12.3 | 18 | 2.0 | 362 | 9.9 | |
| Acute decompensation of heart failure | 317 | 11.4 | 108 | 12.3 | 425 | 11.6 | |
| Other respiratory/pulmonary disease | 170 | 6.1 | 209 | 23.8 | 379 | 10.3 | |
| Arrhythmia | 140 | 5.0 | 14 | 1.6 | 154 | 4.2 | |
| COVID-19† | 135 | 4.8 | 0 | 0 | 135 | 3.7 | |
| AE Bronchitis | 116 | 4.2 | 1 | 0.1 | 117 | 3.2 | |
| Exacerbation of other underlying lung disease | 104 | 3.7 | 14 | 1.6 | 118 | 3.2 | |
| Empyema/lung abscess | 5 | 0.2 | 4 | 0.5 | 9 | 0.2 | |
| Other non-respiratory/pulmonary disease | 450 | 16.1 | 155 | 17.6 | 605 | 16.5 | |
| Number of Comorbidities | | | | | | | |
| Mean (SD) | 3.3 | 2 | 2.8 | 1.7 | 3.2 | 2 | <.0001 |
| Median (IQR) | 3 | 2.0–5.0 | 3 | 1.0–4.0 | 3 | 2.0–4.0 | |
| Duration of hospitalization | | | | | | | |
| Mean (SD) | 4.9 | 4.8 | 9.5 | 11.7 | 6 | 7.4 | <.0001 |

| | | | | | | | |
|---|---|---|---|---|---|---|---|
| Median (IQR) | 3 | 2.0–6.0 | 6 | 3.0–11.0 | 4 | 2.0–7.0 | |
| Visit 1 Discharge Summary | | | | | | | |
| Discharge Disposition | | | | | | | |
| Home without home health care | 2027 | 72.7 | 503 | 57.2 | 2530 | 69 | <.0001 |
| Home with home health care | 441 | 15.8 | 130 | 14.8 | 571 | 15.6 | |
| Transferred to another acute hospital | 23 | 0.8 | 13 | 1.5 | 36 | 1 | |
| Transferred to skilled nursing facility/other rehabilitation facility | 229 | 8.2 | 123 | 14.0 | 352 | 9.6 | |
| Remained in hospital at day 60 | 0 | 0 | 3 | 0.3 | 3 | 0.1 | |
| In-hospital death | 54 | 1.9 | 54 | 6.1 | 108 | 2.9 | |
| Other | 16 | 0.6 | 53 | 6.0 | 69 | 1.9 | |

[a]P-values compare US to Canada and were generated for select variables.

[b]COVID-19 was not included on the list of clinical diagnoses in Canada

,

**Supplementary Table 3. Saliva specimens by collection method**

| **Saliva sample type** | **US** | **Canada** | **All Sites** |
|---|---|---|---|
| Neat Saliva | 1812 (65.91%) | 567 (67.99%) | 2379 (66.40%) |
| Saline Mouth Wash | 541 (19.68%) | 267 (32.01%) | 808 (22.55%) |
| Unknown | 396 (14.41%) | 0 (0.00%) | 396 (11.05%) |
| Total | 2749 | 834 | 3583 |

**Supplementary Table 4: Specimen collection and RSV test results for standard of care or scavenged specimen testing**

| | US | | Canada | | All Sites | |
|---|---|---|---|---|---|---|
| **SOC/scavenged Specimen Collection** | **N** | **%** | **N** | **%** | **N** | **%** |
| Overall | 2790 | 100 | 879 | 100 | 3669 | 100 |
| Specimen Collection and Test Results | | | | | | |
| NPS | 1654 | | 839 | | 2493 | |
| Positive | 106 | 6.4 | 51 | 6.1 | 157 | 6.3 |
| Negative | 1548 | 93.6 | 788 | 93.9 | 2336 | 93.7 |
| Sputum | 9 | | 18 | | 27 | |
| Positive | 1 | 11.1 | 4 | 22.2 | 5 | 18.5 |
| Negative | 8 | 88.9 | 14 | 77.8 | 22 | 81.5 |
| BAL | 21 | | 12 | | 33 | |
| Positive | 1 | 4.8 | 1 | 8.3 | 2 | 6.1 |
| Negative | 20 | 95.2 | 11 | 91.7 | 31 | 93.9 |
| Throat Swab | 0 | | 5 | | 5 | |
| Positive | 0 | N/A | 0 | 0 | 0 | 0 |
| Negative | 0 | N/A | 5 | 100 | 5 | 100 |
| Tracheal Aspirate | 1 | | 0 | | 1 | |
| Positive | 0 | 0 | 0 | N/A | 0 | 0 |
| Negative | 1 | 100 | 0 | N/A | 1 | 100 |
| Pleural Fluid | 0 | | 24 | | 24 | |
| Positive | 0 | N/A | 0 | 0 | 0 | 0 |
| Negative | 0 | N/A | 24 | 100 | 24 | 100 |
| Nasal Wash | 7 | | 0 | | 7 | |

| | | | | | | |
|---|---|---|---|---|---|---|
| Positive | 0 | 0 | 0 | N/A | 0 | 0 |
| Negative | 7 | 100 | 0 | N/A | 7 | 100 |
| Nasal Swab | 0 | | 5 | | 5 | |
| Positive | 0 | N/A | 0 | 0 | 0 | 0 |
| Negative | 0 | N/A | 5 | 100 | 5 | 100 |
| Oropharyngeal Swab | 0 | | 1 | | 1 | |
| Positive | 0 | N/A | 0 | 0 | 0 | 0 |
| Negative | 0 | N/A | 1 | 100 | 1 | 100 |

BAL, bronchoalveolar lavage; NPS, nasopharyngeal swab; SOC, standard of care

**Supplementary Table 5: Increase in RSV detection associated with using additional specimen type results beyond NP swab for all participants in the study (N=3669)**

| **Specimen combinations** | **At least one listed specimen positive for RSV N=3669 N, (%)** | **% increase in RSV detection for combination compared to NPS alone N (95% CI)** |
|---|---|---|
| NPS only | 120 (3.27) | Reference |
| NPS with one other specimen type | | |
| + sputum | 152 (4.14) | 27 (17–38%) |
| + serum | 175 (4.77) | 46 (32–61%) |
| + saliva | 187 (5.10) | 56 (40–73%) |
| NPS with two other specimen types | | |
| + sputum and serum | 203 (5.53) | 69 (51–90%) |
| + saliva and sputum | 206 (5.61) | 72 (53–93%) |
| + saliva and serum | 235 (6.41) | 96 (73–122%) |
| NPS with all 3 other specimen types | 254 (6.92) | 112 (86–141%) |

CI, confidence intervals; N, total number of patients; NPS, nasopharyngeal swab

**Supplementary Table 6: Increase in RSV detection associated with using additional specimen type results beyond NP swab for different populations with specific specimen results available (N variable per specimen type combination)**

| Groups by available specimen combinations | N | Count of participants by NPS positive | Detection rate with NPS positive (per 100 participants) | Count of participants by any listed specimen positive | Detection rate with any listed specimen positive (per 100 participants) | % Increase in RSV detection, 95%CI (Any listed/NPS) |
|---|---|---|---|---|---|---|
| All four specimens | 417 | 24 | 5.76 | 52 | 12.47 | 117 (62–191%) |
| NPS with two other specimen types | | | | | | |
| +Saliva and Sputum | 1162 | 64 | 5.51 | 109 | 9.38 | 70 (46–99%) |
| +Saliva and Serum | 1179 | 48 | 4.07 | 113 | 9.58 | 135 (90–192%) |
| +Sputum and Serum | 427 | 24 | 5.62 | 51 | 11.94 | 113 (59–184%) |
| NPS with one other specimen type | | | | | | |
| +Saliva | 3583 | 118 | 3.29 | 185 | 5.16 | 57 (41–75%) |
| +Sputum | 1212 | 66 | 5.45 | 98 | 8.09 | 48 (29–70%) |
| +Serum | 1225 | 48 | 3.92 | 103 | 8.41 | 115 (75–164%) |

CI, confidence intervals; N, total number of patients; n, number of RSV-positive patients; NPS, nasopharyngeal swab

**Supplementary Table 7: Characteristics of RSV participants identified by NPS compared with RSV participants identified by other specimen types only**

| | **NPS Positive**[a] | | **NPS Negative & Saliva Positive** | | **NPS Negative & Sputum Positive** | | **NPS Negative & Serology Positive** | | **NPS Negative & Saliva/Sputum/Serology Positive** | |
|---|---|---|---|---|---|---|---|---|---|---|
| | **N** | **%** | **N** | **%** | **N** | **%** | **N** | **%** | **N** | **%** |
| Overall | 120 | 100 | 67 | 100 | 32 | 100 | 55 | 100 | 134 | 100 |
| Age in Years | | | | | | | | | | |
| Median (IQR) | 70.5 | 60.0–81.0 | 70 | 59.0–83.0 | 67 | 55.0–76.5 | 64 | 54.0–75.0 | 66.5 | 57.0–78.0 |
| 40-64 | 44 | 36.7 | 27 | 40.3 | 14 | 43.8 | 28 | 50.9 | 62 | 46.3 |
| ≥65 | 76 | 63.3 | 40 | 59.7 | 18 | 56.3 | 27 | 49.1 | 72 | 53.7 |
| Time from symptom onset to specimen collection in days[b] | | | | | | | | | | |
| Median (IQR) | 5 | 3.0–6.0 | 4 | 2.0–7.0 | 3 | 2.0–6.0 | 5 | 3.0–9.0 | 4 | 2.0–8.0 |
| 0 days (on the same day) | 1 | 0.8 | 2 | 3 | 0 | 0 | 0 | 0 | 2 | 1.5 |
| 1-2 days | 26 | 21.7 | 20 | 29.9 | 13 | 40.6 | 12 | 21.8 | 38 | 28.4 |
| 3-4 days | 32 | 26.7 | 17 | 25.4 | 7 | 21.9 | 12 | 21.8 | 31 | 23.1 |
| ≥5 days | 61 | 50.8 | 28 | 41.8 | 12 | 37.5 | 31 | 56.4 | 63 | 47 |
| Signs and Symptoms | | | | | | | | | | |

| | | | | | | | | | | |
|---|---|---|---|---|---|---|---|---|---|---|
| Dyspnea (shortness of breath/air) | 98 | 81.7 | 53 | 79.1 | 26 | 81.3 | 44 | 80 | 108 | 80.6 |
| Cough | 97 | 80.8 | 49 | 73.1 | 29 | 90.6 | 30 | 54.5 | 89 | 66.4 |
| Sputum production | 58 | 48.3 | 28 | 41.8 | 26 | 81.3 | 16 | 29.1 | 54 | 40.3 |
| Wheezing | 44 | 36.7 | 18 | 26.9 | 12 | 37.5 | 7 | 12.7 | 28 | 20.9 |
| Hypoxemia (low blood oxygen) | 30 | 25 | 9 | 13.4 | 4 | 12.5 | 9 | 16.4 | 17 | 12.7 |
| Nasal congestion | 20 | 16.7 | 11 | 16.4 | 6 | 18.8 | 5 | 9.1 | 18 | 13.4 |
| Rhinorrhea (Runny nose) | 20 | 16.7 | 13 | 19.4 | 7 | 21.9 | 5 | 9.1 | 19 | 14.2 |
| Sore throat | 15 | 12.5 | 3 | 4.5 | 2 | 6.3 | 2 | 3.6 | 7 | 5.2 |
| Hoarseness | 7 | 5.8 | 2 | 3 | 0 | 0 | 0 | 0 | 2 | 1.5 |
| Final Clinical Diagnosis | | | | | | | | | | |
| Community-acquired pneumonia | 37 | 30.8 | 20 | 29.9 | 7 | 21.9 | 9 | 16.4 | 34 | 25.4 |
| Acute exacerbation of COPD | 28 | 23.3 | 10 | 14.9 | 10 | 31.3 | 6 | 10.9 | 21 | 15.7 |
| Acute bronchitis | 18 | 15 | 5 | 7.5 | 2 | 6.3 | 2 | 3.6 | 7 | 5.2 |
| Other respiratory/pulmonary disease | 14 | 11.7 | 6 | 9 | 3 | 9.4 | 7 | 12.7 | 14 | 10.4 |

| | | | | | | | | | |
|---|---|---|---|---|---|---|---|---|---|
| Acute decompensation of heart failure | 6 | 5 | 8 | 11.9 | 1 | 3.1 | 7 | 12.7 | 16 | 11.9 |
| Arrhythmia | 5 | 4.2 | 2 | 3 | 2 | 6.3 | 2 | 3.6 | 5 | 3.7 |
| Exacerbation of other underlying lung disease | 4 | 3.3 | 3 | 4.5 | 4 | 12.5 | 2 | 3.6 | 6 | 4.5 |
| Acute chest pain/cardiac ischemia | 1 | 0.8 | 2 | 3 | 0 | 0 | 5 | 9.1 | 7 | 5.2 |
| Other non-respiratory/pulmonary disease | 6 | 5 | 8 | 11.9 | 3 | 9.4 | 14 | 25.5 | 20 | 14.9 |
| Immunocompromised | 41 | 34.2 | 25 | 37.3 | 10 | 31.3 | 21 | 38.2 | 50 | 37.3 |
| Disease Severity | | | | | | | | | | |
| ICU | 9 | 7.5 | 4 | 6 | 3 | 9.4 | 6 | 10.9 | 11 | 8.2 |
| Mechanical ventilation | 6 | 5 | 1 | 1.5 | 1 | 3.1 | 1 | 1.8 | 3 | 2.2 |
| ICU or mechanical ventilation | 9 | 7.5 | 5 | 7.5 | 3 | 9.4 | 6 | 10.9 | 12 | 9 |
| Not severe | 111 | 92.5 | 62 | 92.5 | 29 | 90.6 | 49 | 89.1 | 122 | 91 |
| Death | 2 | 1.7 | 1 | 1.5 | 1 | 3.1 | 0 | 0 | 2 | 1.5 |

COPD, chronic obstructive pulmonary disorder; ICU, intensive care unit; NPS, nasopharyngeal swab

[a]NP swab test positive may include participants who are also saliva or sputum or serology test positive.

[b]Some participants may have symptom onset date missing.
